# Paper 3: - Towards a Knowledge Sharing Maturity Model for medical imaging departments

**DOI:** 10.1101/2024.04.01.24305015

**Authors:** Maryam Almashmoum, James Cunningham, John Ainsworth

**Affiliations:** Division of Informatics Imaging and Data Sciences, School of Health Sciences Faculty of Biology, Medicine, and Health The University of Manchester, Manchester Academic Health Science Centre, Manchester, United Kingdom; Nuclear Medicine Department, Faisal Sultan Bin Eissa, Kuwait Cancer Control Center, Kuwait

**Keywords:** Knowledge management, knowledge sharing, medical imaging departments, cancer centres, the Christie, KCCC, maturity model, factors, indicators, measurement

## Abstract

**Background:** Knowledge sharing in medical imaging departments is being driven by the need to improve healthcare services, develop healthcare professionals’ skills, and reduce repetitive mistakes. It is considered an important step in the implementation of knowledge management solutions. By following a maturity model of knowledge sharing, knowledge sharing practices can be improved. The aim of this study was to develop a maturity model knowledge sharing in medical imaging department for helping managers to assess the level of maturity for knowledge sharing practices.

**Objectives:** In the modern healthcare institutions, improvements in healthcare professionals’ skills and healthcare services are often driven through practicing knowledge sharing behaviours. To understand the level of maturity of knowledge sharing, mangers can follow the indicators of maturity model knowledge sharing and its measurements in order to identify the current level and move to the next level.

**Methods:** This study was conducted in three stages: An ‘overview stage’ which highlighted the factors that affect knowledge sharing practices in medical imaging departments; an ‘Analysis factor stage’ which was designed to assess the factors that affect knowledge sharing by using a concurrent mixed method approach’s (questionnaires, and semi-structured interviews) in two medical imaging departments; and ‘Structuring maturity model knowledge sharing stage’, where a maturity model of knowledge sharing was developed based on the findings of the other stages.

**Results:** The model presented in this study includes 17 indicators divided into 11 components. Those components derived from the findings of the questionnaires and semi-structured interviews that were applied in the medical imaging departments. It consists of five maturity levels: initial, aware, define, managed, and optimised. In each level were included measurements in order to help managers to assess the current level by answering to the measurement’s questions.

**Conclusion:** This maturity model of knowledge sharing in medical imaging departments allows managers and policy makers to measure the maturity level of knowledge sharing in those departments. Although the model has been applied to the medical imaging departments, it might easily be modified to apply it to other institutions.

## Introduction

Knowledge is an essential asset in achieving successful institutions practices, and sharing knowledge gives a sustainable competitive advantage to modern institutions [1] [2]. Accessing knowledge is the main step in problem solving and decision making [3]. Knowledge is a mixture of experiences, thoughts, ideas, values, and information. In the context of institutions, knowledge can be shared and transformed among employees within institutions to create new experiences and information that did not exist before [4]. Healthcare institutions, for example, hospitals, and specialised centres have to apply knowledge management (KM) system in order to build a proper and effective network among all healthcare providers [4]. The reason for implementing knowledge management system is mainly because of its complexity, and the huge number of knowledge-based resources that need to be managed [5]. Implementing knowledge management depends on understanding its processes (knowledge creation, knowledge capture, knowledge sharing, and knowledge applications). Moreover, identifying a clear framework to follow is vital to target any weaknesses in each step, and creating a good environment for healthcare professionals. As a result, health care services, and patient outcomes can be improved [5].

Knowledge sharing is considered an important factor to improve health care services, and an essential step for successful knowledge management within institutions as it can lead to improve healthcare settings performance, save them time, reduce costs, and increase health education levels [6]. It plays an important role in providing great accountability and establishing good practices in health planning and policymaking. Abzari et al. [7] stated that knowledge sharing is considered an essential factor in successful institutional performance. It refers to the act of sharing both tacit and explicit knowledge, such as thoughts, ideas, and experiences from one person, group, or institution to another in order to formulate new knowledge [8]. Knowledge sharing is one of the challenges for healthcare institutions due to the variety of resources and knowledge. Healthcare institutions are complex environments given the variety of specialities in each department and who operates institutional resources in each department [9].

Medical imaging departments are essential in any healthcare institution due to the importance of performing important procedures for the pre-treatment plan and interpreting results. Based on the observation at the cancer centre, there are several problems facing knowledge sharing practices in the medical imaging department such as a lack of awareness of the importance of knowledge sharing, difficulty sharing knowledge among healthcare professionals, and a lack of implementation of information and communication technology (ICT) [10]. Creating a communication environment is important to control the amount of knowledge shared among healthcare professionals in their institutions [11]. Additionally, dealing with healthcare institutional issues is directly related to human capital [12]. Sharing knowledge among healthcare professionals is critical to apply that knowledge in their daily work and allow them to create new knowledge that is needed for developing the institutional process. Both tacit and explicit knowledge are important to enhance the knowledge sharing process among healthcare professionals because they are directly related to their experience, and skills [13]. Therefore, identifying a clear maturity model for knowledge sharing in medical imaging is important to help policymaker, and leaders follow it to enhance knowledge sharing practices among healthcare professionals.

## Literature review

Maturity in this context refers to the degree to which technology, institutional process or frameworks evolve over time [14, 15]. In institutions, a maturity model (MM) can be methodically used to define operations and identify stages which can lead to policy plans [16]. The concept of the maturity model was developed in the early 1970s [17]. It is increasingly applied in the field of information systems [18] and is established in many fields such as knowledge management, information management, software performance and management [19–22]. The purpose of maturity models is to provide a clear model based on an institution’s capabilities in a certain managerial area by using a set of criteria and related evaluation methodologies [23]. It can be a powerful tool that helps identify strengths and weaknesses in a specific area [24]. Bititci et al., [25] indicated that maturity models have a positive impact on improving institutional performance and respond to many challenges by describing each step and stage properly.

Healthcare institutions face challenges in achieving best practices in knowledge management implementation [26]. Maturity in knowledge management is defined as the degree to which knowledge assets are effectively managed and controlled within institutions [27]. There have been several maturity models for evaluating and descripting these areas of management that have been proposed [14–16, 28–30]. Some other applications of maturity models have been applied to healthcare intuition information systems [31].

There is a continuing need to structure and develop a new Knowledge Sharing Maturity Model (KSMM) for enhancing knowledge sharing practices among employees in the medical imaging departments [32]. KSMM helps decision-makers to achieve institutional tasks and improve the quality of patient outcomes [33]. There are several models of knowledge sharing [34–37]. Despite the variety of these models, few of them focus on knowledge sharing in healthcare in general and in the medical imaging department in particular [38]. There are several factors that affect knowledge sharing among healthcare professionals, including the maturity model of multidisciplinary team meetings, community of practice information and communication technology, social media networks, PACS, tele-medicine, and digital library [39–45]. Creating a maturity model for knowledge sharing that consists of all factors that affect their behaviours is vital to implementing a knowledge sharing environment among healthcare professionals.

To date, there is no maturity model to assess knowledge sharing practices among healthcare professionals in general hospitals as a whole, or in medical imaging departments specifically. Developing a maturity model to evaluate the level of maturity of knowledge sharing behaviours is a challenge due to the interactions of healthcare professionals’ behaviours with each other on one hand and with ICT on the other, and is dependent on human beings, cultural environments and technological facilitators [46].

## Aim and Research questions

The aim of this paper is to structure and present the steps of the development of a KSMM for understanding how healthcare professionals share knowledge, and to define the proper stages of achieving the best knowledge practices among healthcare professionals in medical imaging departments. Therefore, the key research questions are: (1) What are the main components that structured the KSMM? (2) What are the stages of MM that help managers in medical imaging departments need to follow in order to evaluate and enhance knowledge sharing practices among healthcare professionals? (3) What are the indicators that control the KSMM, and how can managers measure them.

## Research objectives

This paper has various objectives. Firstly, to identify the main stages and components that relate to KSMM. Secondly, the main characteristics of each stage involved in the MM explained in depth. Finally, to create a proposal for a MM that includes the knowledge sharing influencing indicators that affect knowledge sharing practices among healthcare professionals in medical imaging departments. Additionally, the measurements for each indicator that form the components of the KSMM will be presented.

## Research methodology

This paper illustrates the development of a maturity model for knowledge sharing in medical imaging departments. There were three stages conducted in this study, as shown in Figure 1. First an “overview stage”, consisting of a review of previous literature that identified factors that affect knowledge sharing practices in healthcare institutions in general and medical imaging departments specifically. Second the “analysis factors” stage, which consists of applying several methodologies to examine those factors. Third the “structuring” stage. Here, the initial maturity model for knowledge sharing in the medical imaging department was be developed.

**Figure 1:**
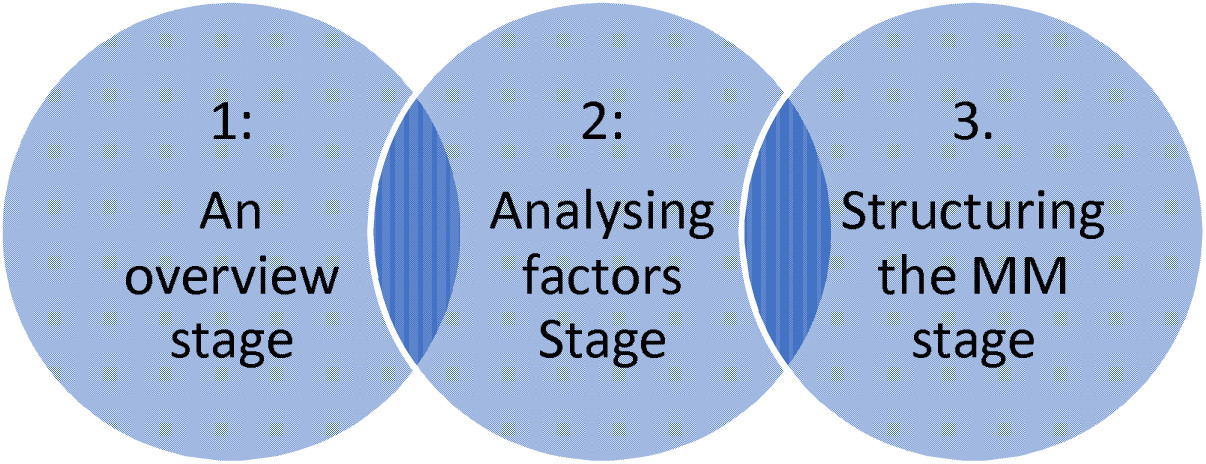
The three stages conducted in this study.

## An overview stage

An overview stage illustrated the factors that affect knowledge sharing in medical imaging departments at the general hospital and cancer centres specifically. There were several studies that identified knowledge sharing factors were divided into three categories: individual factors, administrative factors, and technological factors [38, 47–59]. Any factor that has a positive impact on enhancing knowledge sharing is called a facilitator. On the other hand, any factors that hinder knowledge sharing practices is called a barrier. Based on previous literature, the factors that affect knowledge sharing in medical imaging departments are the same whether in a general hospital or a cancer centre. However, they are different in terms of terminology because the nature of the cancer centre is mainly concerned with treating cancer cases, which need more than one specialty to end up with an appropriate protocol to treat specific cases [60]. Therefore, documenting the factors that affect knowledge sharing in medical imaging departments from the previous literature is important in order to test those factors in the next stage [60]. Those factors are shown in Table 1.

**Table 1:** Factors that affect knowledge sharing in the medical imaging departments in general hospitals Vs cancer centres [60].

| **Types of factors** | **Facilitators terminologies in medical imaging departments in general hospitals** | **Facilitators terminologies in medical imaging departments in cancer centres** |
| --- | --- | --- |
| Individual factors | -Trust<br>-Positive attitudes<br>-Awareness<br>-Experience<br>-Intrinsic Motivation<br>-Personality<br>-Self-esteem<br>-Self-efficacy | -Trust<br>-Positive attitudes<br>-Awareness<br>-Experience<br>-Intrinsic Motivation<br>-Personality<br>-Self-esteem<br>-Self-efficacy |
| Departmental factors | -Communities of Practice (CoP), inter-professional collaboration<br>-leadership<br>-Culture<br>-Teamwork<br>-Extrinsic motivation<br>-Learning and training<br>-Doctor rounds<br>-Departmental arrangements | -Multi disciplinary team (MDT), Community of oncologists<br>-leadership<br>-Culture<br>-Teamwork<br>-Extrinsic motivation<br>-Learning and training<br>-Doctor rounds<br>-Departmental arrangements |
| Technological factors | -ICT (PACS, social media, intranet, extranet, tele-medicine, tele-radiology)<br>-Network<br>-Digital library | -ICT (PACS, social media, intranet, extranet, tele-medicine, teleradiology)<br>-Network<br>-Digital library |

## Analysis factors stage

In this stage, to examine and analyse those factors, mixed methods are used in this study. The mix method consists of questionnaires, and semi-structured interviews [61]. A triangulation – validate mixed methods design was used in this study. The questionnaire was structured based on the previous studies, and the semi-structured interview questions were formed based on the factors in the previous stage in order to examining those factors through distributing questionnaires and conducting semi-structured interview among healthcare professionals who are working in the medical imaging department at two cancer centres [62–64]. The questionnaire was divided into three parts: the demographic section, knowledge sharing practices, and factors that affect knowledge sharing practices in the medical imaging departments. The questionnaires were distributed electronically using the Qualtrics survey tool. The semi-structured interviews were conducted via Microsoft Teams. Those methods were applied among healthcare professionals in the medical imaging department of two cancer centres: the Christie hospital and Kuwait cancer Control Centre.

## Structuring MM stage

In this stage a maturity model for knowledge sharing was developed. Based on the identifying factors that affect knowledge sharing practices in medical imaging departments, a clear vision was developed in order to build a KSMM in a proper way, helping the managers and policymakers to follow those steps and understand how those factors are related to each other in different stages. Furthermore, it helps them either to implement knowledge sharing environment, or to identify the weak points of knowledge sharing practices.

## Results of mixed methods

In this paper, we used a triangulation concurrent mixed method approach. An electronic survey was distributed among healthcare professional in two cancer centres from February 2023 until July 2023. It consists of 65 questions divided into three sections. To assess the factors that affect knowledge sharing practices, the Likert scale with seven points was used to evaluate those factors, Multimedia Appendix 1 shown the survey questions. A total of 85 responses were received. Qualtrics XM software was used to analyse the quantitative data. Those factors were divided into three categories based on the systematic review: individual factors, departmental factors, and technological factors [60].

The semi-structured interviews were conducted online using Microsoft Teams among 13 healthcare professionals who are working in the medical imaging departments at two cancer centres. Thematic analysis was used to analyse the qualitative data. Codes were organised by using NVivo software. This was done in order to validate the quantitative methods, understand their views, and understand how knowledge sharing practices are going in those centres. Additionally, to assess if there are a clear policy to adopt knowledge sharing practices. Multimedia appendix 2 shows the interview questions and consent form. All those methods were applied under the ethical improvement programme at both centres.

The results revealed that there are 10 components derived from 5 categories, as shown in Table 1. Those categories start with awareness, which is considered the as main step to adopting knowledge sharing practices in the department. Awareness of the importance of knowledge sharing in developing healthcare professionals’ skills and improving healthcare services is important to increase their willingness to share their knowledge. The results of the quantitative method showed that healthcare professionals in both cancer centres have a high level of awareness of the importance of knowledge sharing. The second category is related to the types of knowledge sharing. It is divided into two categories: tacit and explicit knowledge. Tacit knowledge is the most dominant type of knowledge in the medical imaging department because of several meetings, and specialised meetings (multidisciplinary team meetings, Community of practices, and community of oncologists) are occurred in those departments. In contrast, explicit knowledge exits in the endorsed documents, protocols, policies, and procedures manuals. Therefore, understanding both types of knowledge and organising them is important to increase knowledge sharing practices.

The third, fourth, and fifth categories are related to the factors that affect knowledge sharing practices: individual, departmental, and technological factors. The mean scores of the factors fall between Somewhat Agree to Strongly Agree, which reveals that healthcare professionals are believed in the importance of those factors in enhancing knowledge sharing behaviours. The individual factors consist of two components: communication among health professionals, and personality & positive attitudes. Building trust relationships among healthcare professionals is important to allow them to share their experiences. Additionally, most of the respondents agreed that intrinsic motivation has a positive impact on sharing knowledge by increasing their self-efficacy, and self-esteem through giving them opportunities to practice their abilities and experiences. There is a strong relationship between (personality and positive attitudes), and increased knowledge sharing practices. The respondents indicated that knowledge sharing behaviours occur among individuals. Each of them has a specific personality and attitude towards knowledge sharing practices. Some of them like to share their knowledge in large groups, while other feel relief when they share it in small groups.

The departmental factors have a big responsibility to enhance knowledge sharing based on respondents’ thoughts. It consists of five components: leadership and culture, achieving departmental task, continues education, decision making, and infrastructure and workforce. The participants agreed that a leader has a responsibility to create a culture of communication that allows healthcare professionals to practice knowledge sharing behaviours. Developing healthcare professionals’ skills required setting a clear plan for practicing continuing education activities such as attending conferences, lectures, training sessions, and workshops.

Offering them space and time is vital to allowing them to practice knowledge sharing activities through organising tasks among them and giving them empty spaces. To achieve departmental tasks, working within a team is crucial to increase the number of tasks achieved. Making a decision regarding patient treatment is one of the important tasks in the department. That task was created in highly specialised meetings such as: Multidisciplinary teams (MDTs), and communities of practices (CoP).

Technological factors consist of several technological modalities such as PACS, social media, intranet, extranet, tele-medicine, tele radiology that require a high-speed networking. These factors consist of two components: stored and shared electronic data electronically, and access to the electronic databases. The respondents illustrated that using the technology requires skills for using it efficiently. Moreover, accessing the databases is vital to expanding healthcare professionals’ knowledge about up-to date treatment plans used to treat cancer patients.

## Maturity Model development

The development of the maturity model was derived from the systematic review that identified the factors that affect knowledge sharing behaviours in the medical imaging department, and concurrent mixed methods that evaluated those factors in two cancer centres. The non-probability sampling technique was used for mixed methods. Self-selection sampling technique for the quantitative methods, and snowball sampling technique for the qualitative method. The results indicated that KSMM consists of 17 indicators divided into 11 components. Those components represented the five main categories: awareness, knowledge sharing repository, individual factors, departmental factors, and technological factors. Table 2 shows the details of the maturity model for knowledge sharing. Knowledge sharing is a dynamic process among healthcare professionals to manage the institutional process and interaction with information and communication technology infrastructures. As a consequence, those components were incorporated into the five maturity levels. Those level were adopted from the work of the Lee & Kankanhalli [16] that indicated that there are five levels of maturity in knowledge management (initial, aware, defined, managed, and optimized) based on people, process, and technology. The maturity model of knowledge sharing for the medical imaging department presented in Table 2. For each indicator, there are measurement questions that help managers and policymakers to assess each indicator.

**Table 2:** The maturity model of knowledge sharing for the medical imaging departments.

|  |  |  |
| --- | --- | --- |
| Categories | Components | Indicators |
| Awareness | -Awareness and willingness | 1. Awareness and willingness towards KS |
| Types of knowledge sharing<br>-Tacit knowledge<br>-Explicit knowledge | -Understanding and organizing knowledge sharing repository | 2. Structured, and collected both types of knowledge |
| Individual Factors<br>-Trust<br>-Positive attitudes<br>-Experience<br>-Intrinsic Motivation<br>-Personality<br>-Self-esteem<br>-Self-efficacy | - Communication among healthcare professionals.<br>-Personality and positive attitudes | 3. Building trust among healthcare professionals and share their experience<br>4. Increased intrinsic motivation (self-efficacy, and self-esteem)<br>5. Personality and communication among healthcare professionals |
| Departmental factors<br>-Multi disciplinary team (MDT), Community of oncologists<br>-leadership<br>-Culture<br>-Teamwork<br>-Extrinsic motivation<br>-Learning and training<br>-Doctor rounds<br>-Departmental arrangements | -Leadership and culture<br>-Achieving departmental tasks<br>-Continuous education and develop HCP skills<br>-Decision making<br>-Infrastructure and workforce | 6. Structured leadership and creating culture<br>7. Handover policy<br>8. Creating teamwork<br>9. Organising (learning lectures, workshops, training sessions, doctor rounds, and participation in conferences<br>10. Regular meeting<br>11. MDT and Cop making decision<br>12. Meeting room and office layout<br>13. Enhanced extrinsic motivation<br>14. Organised work process |
| Technological factors<br>- ICT (PACS, social media, intranet, extranet, tele-medicine, teleradiology)<br>- Network<br>- Digital library | - Stored and shared patient data electronically<br>- Access to the electronical databases | 15. Strong network<br>16. Implementation information, communication technology, and maintenances.<br>17. Digital library |

**Table 2:**
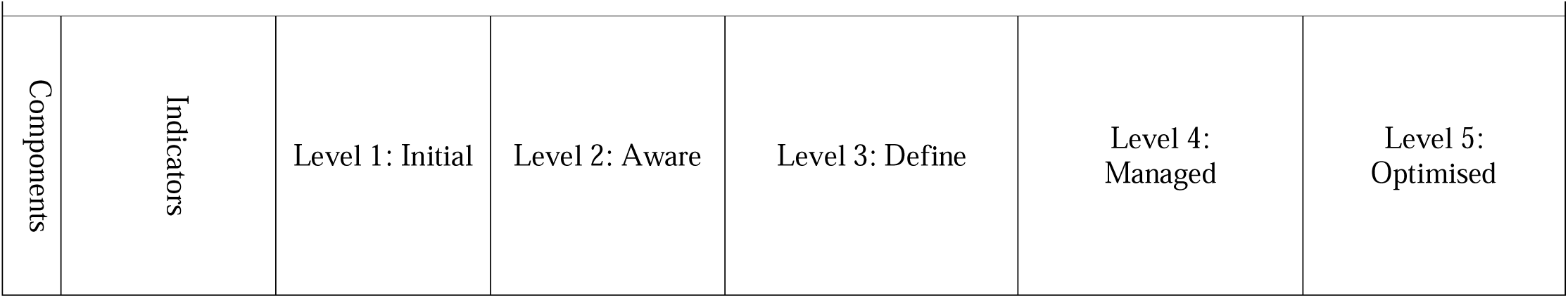
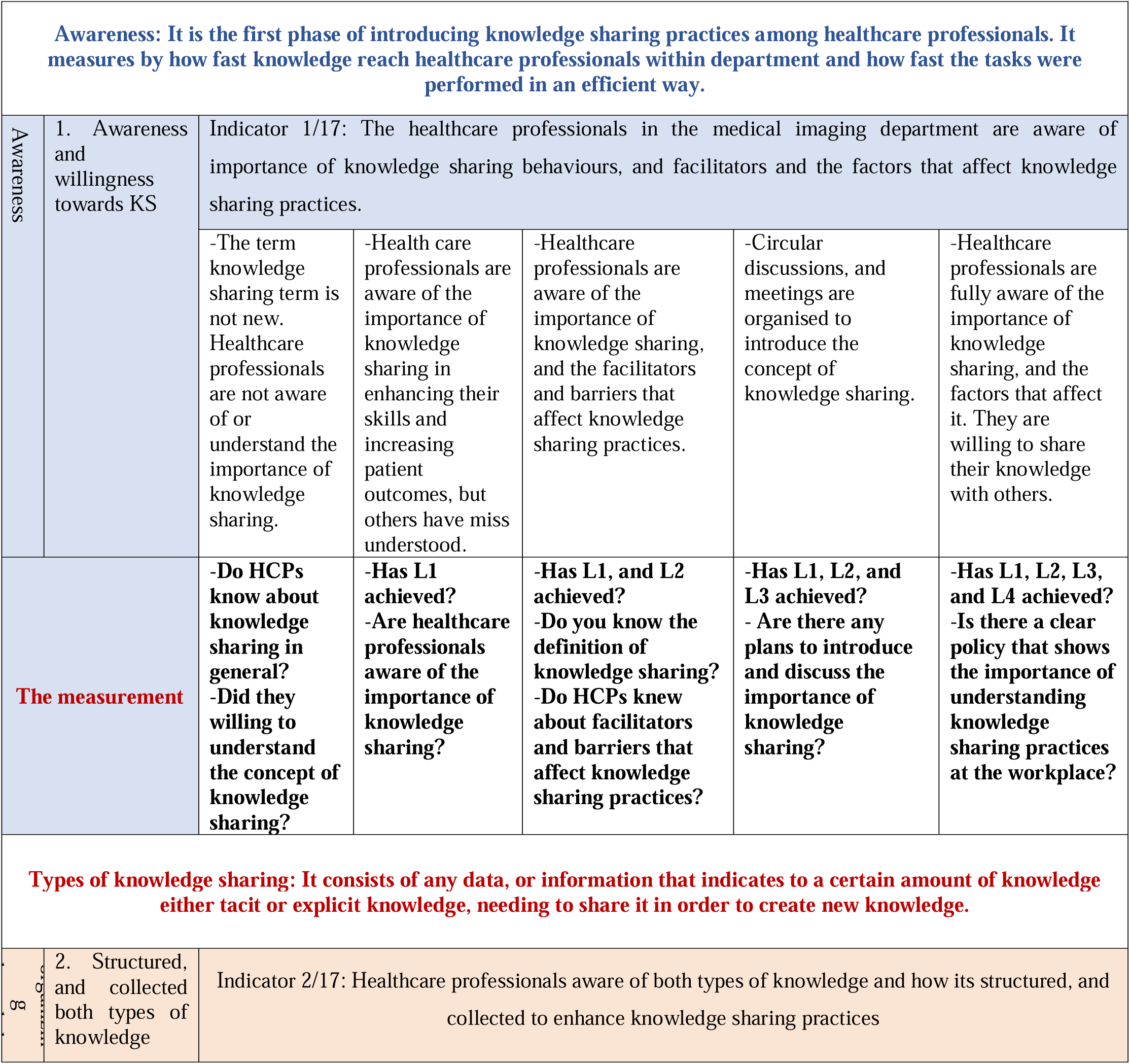
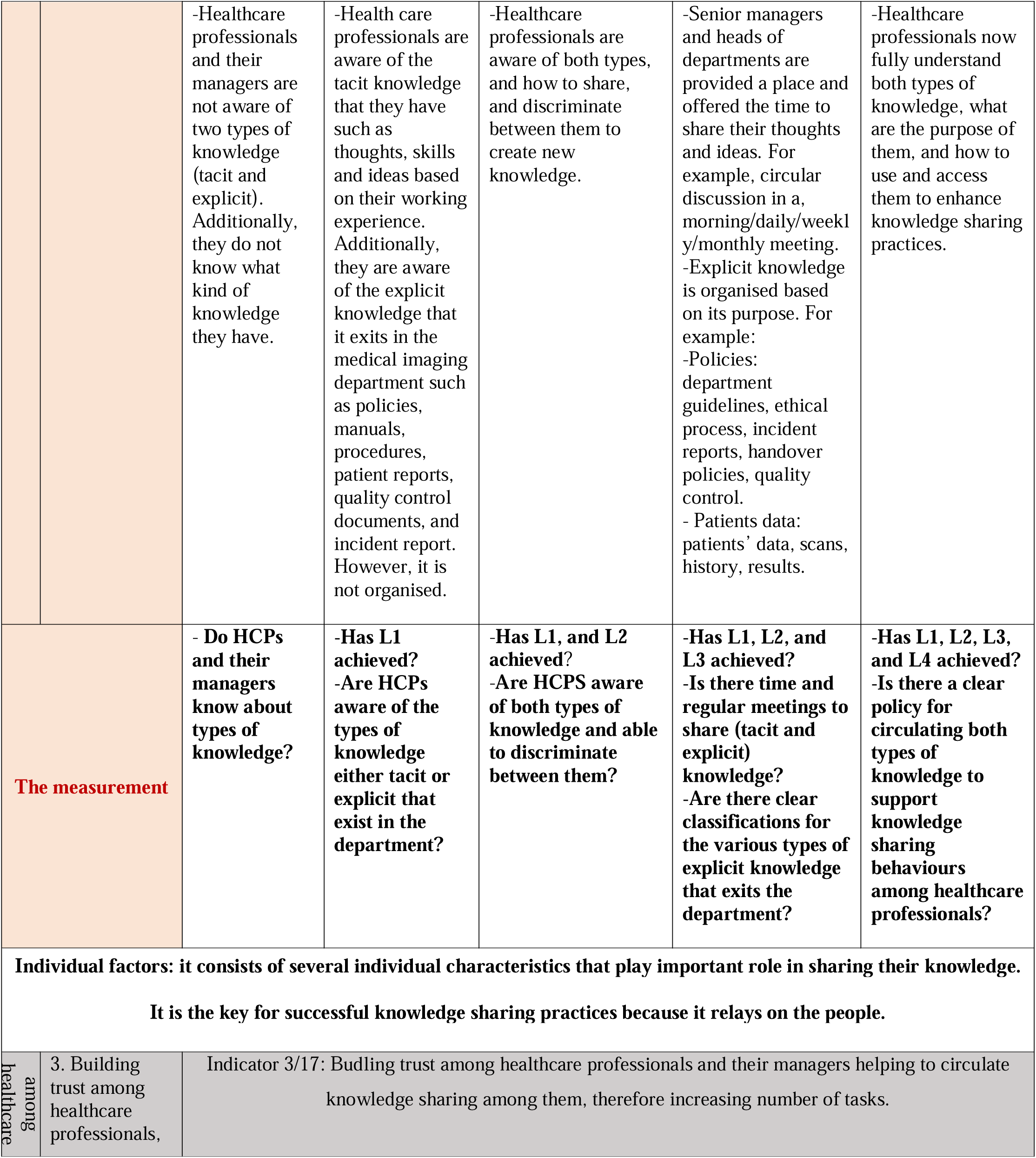
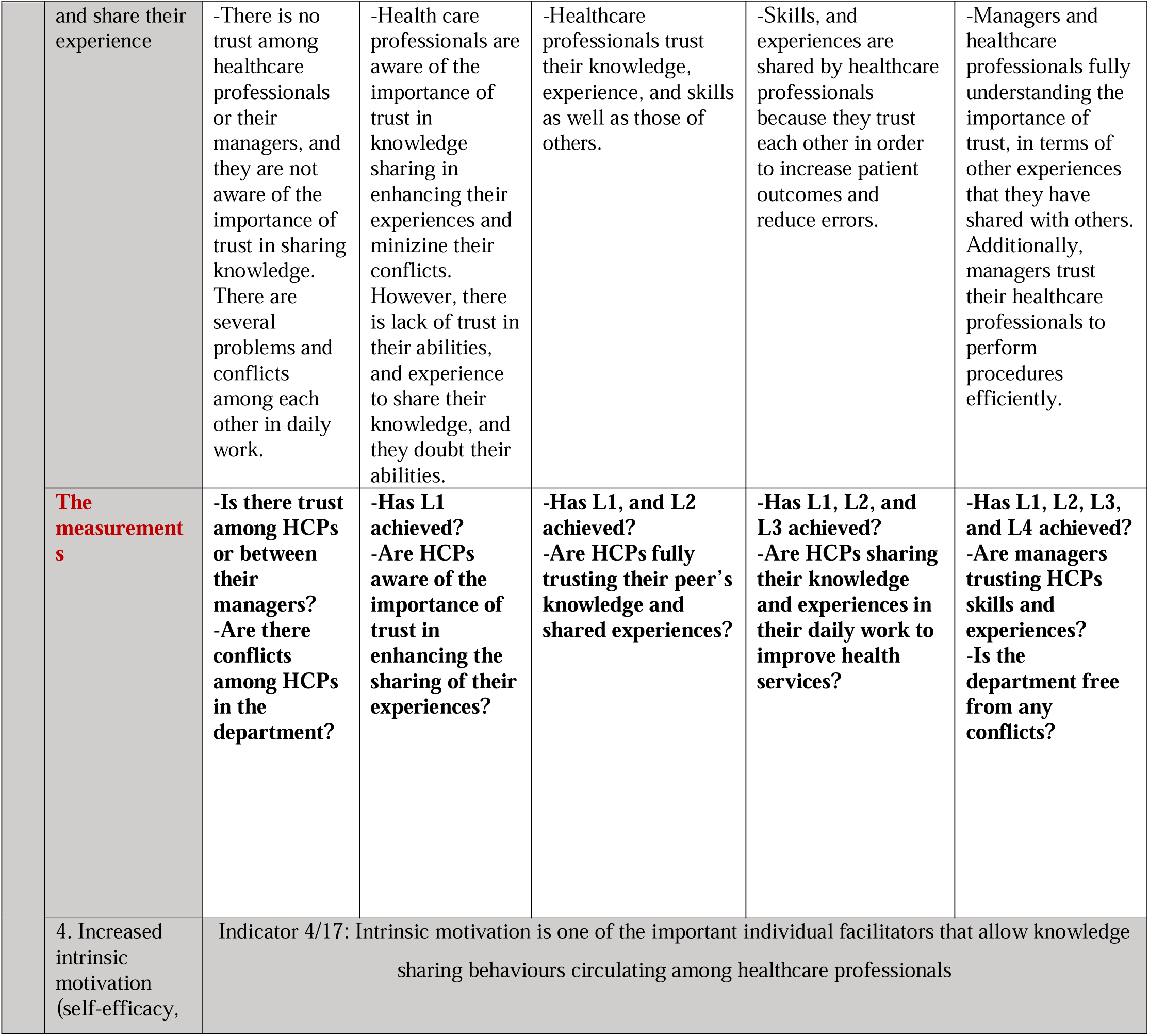
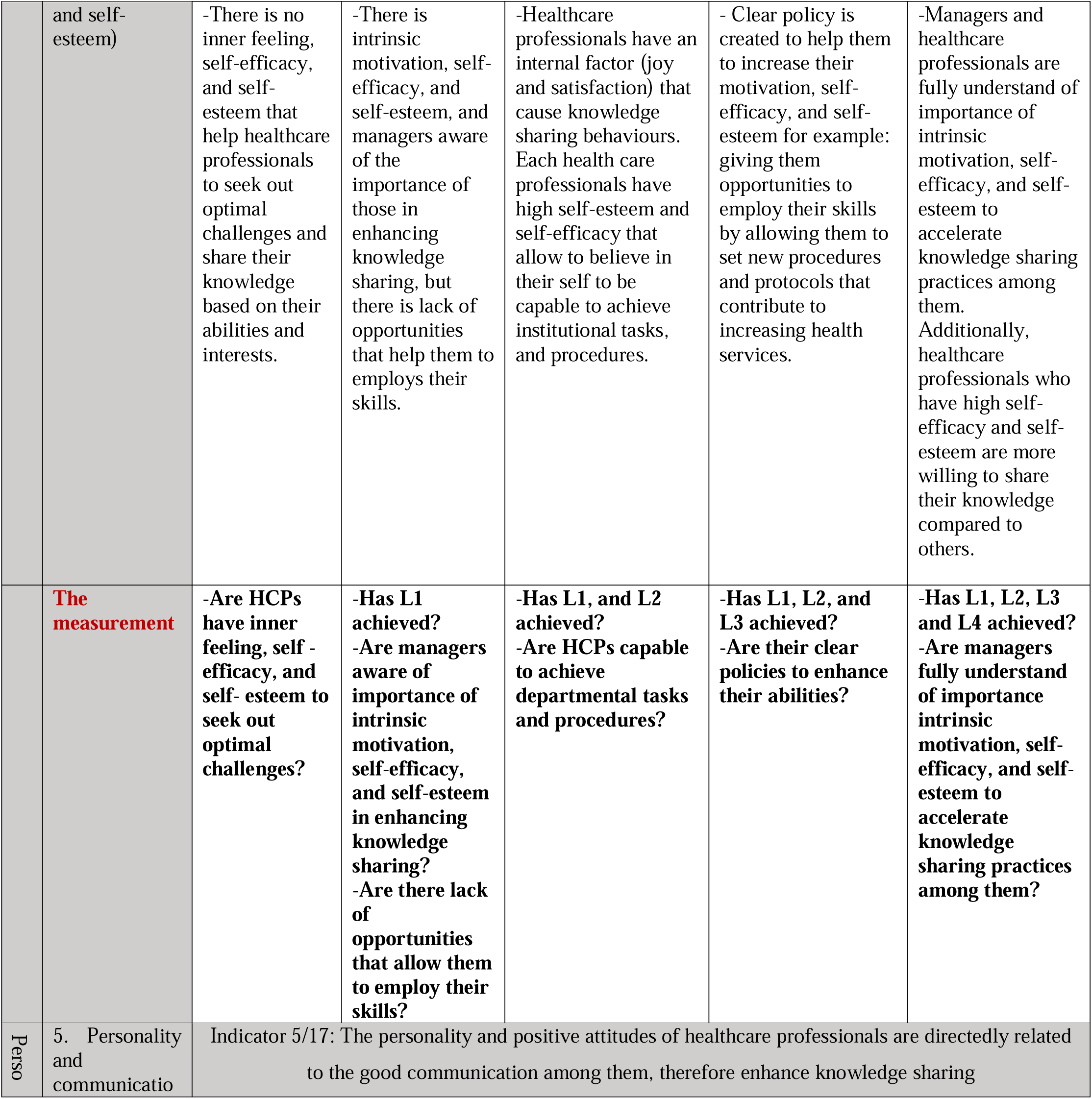
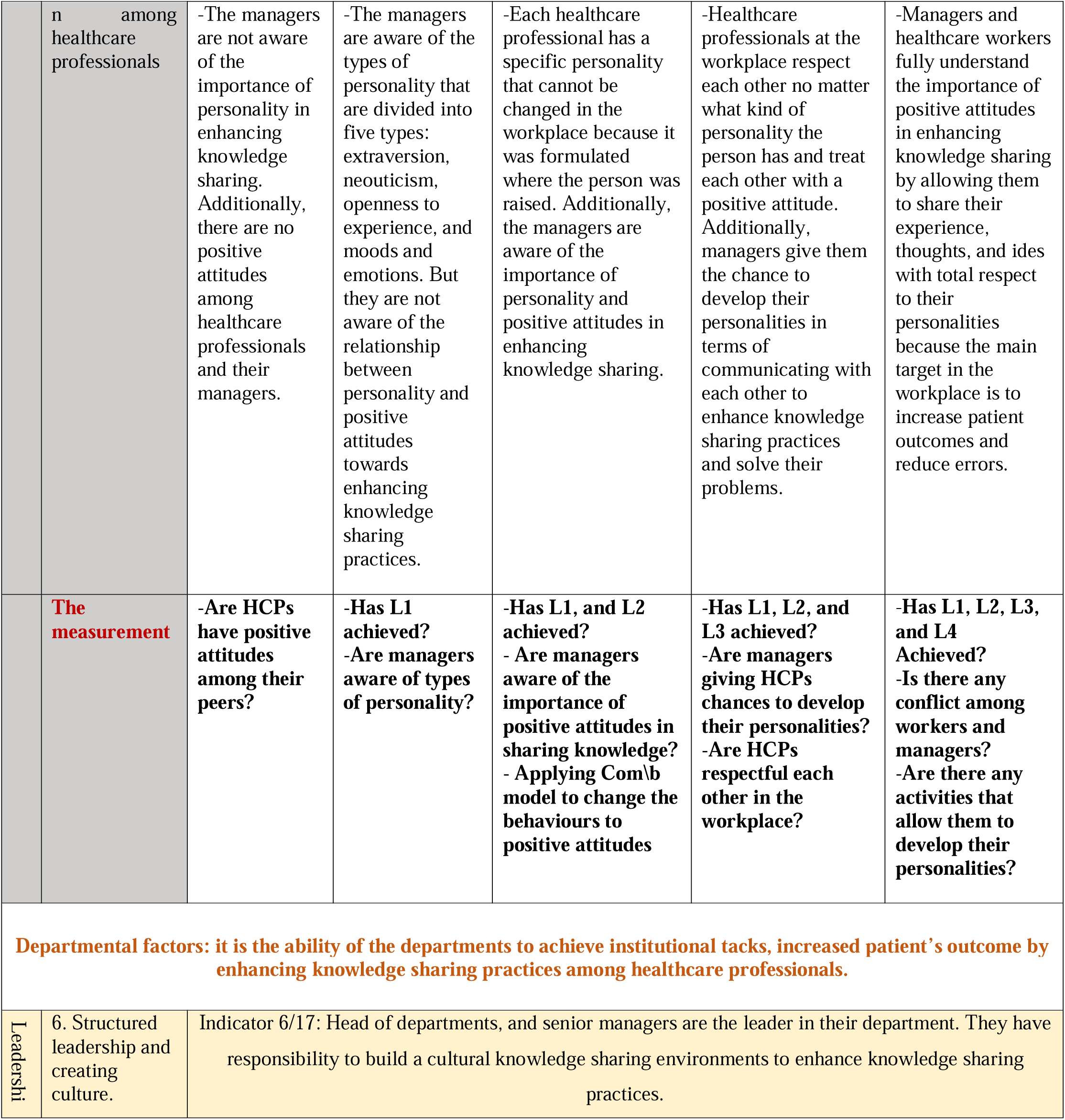
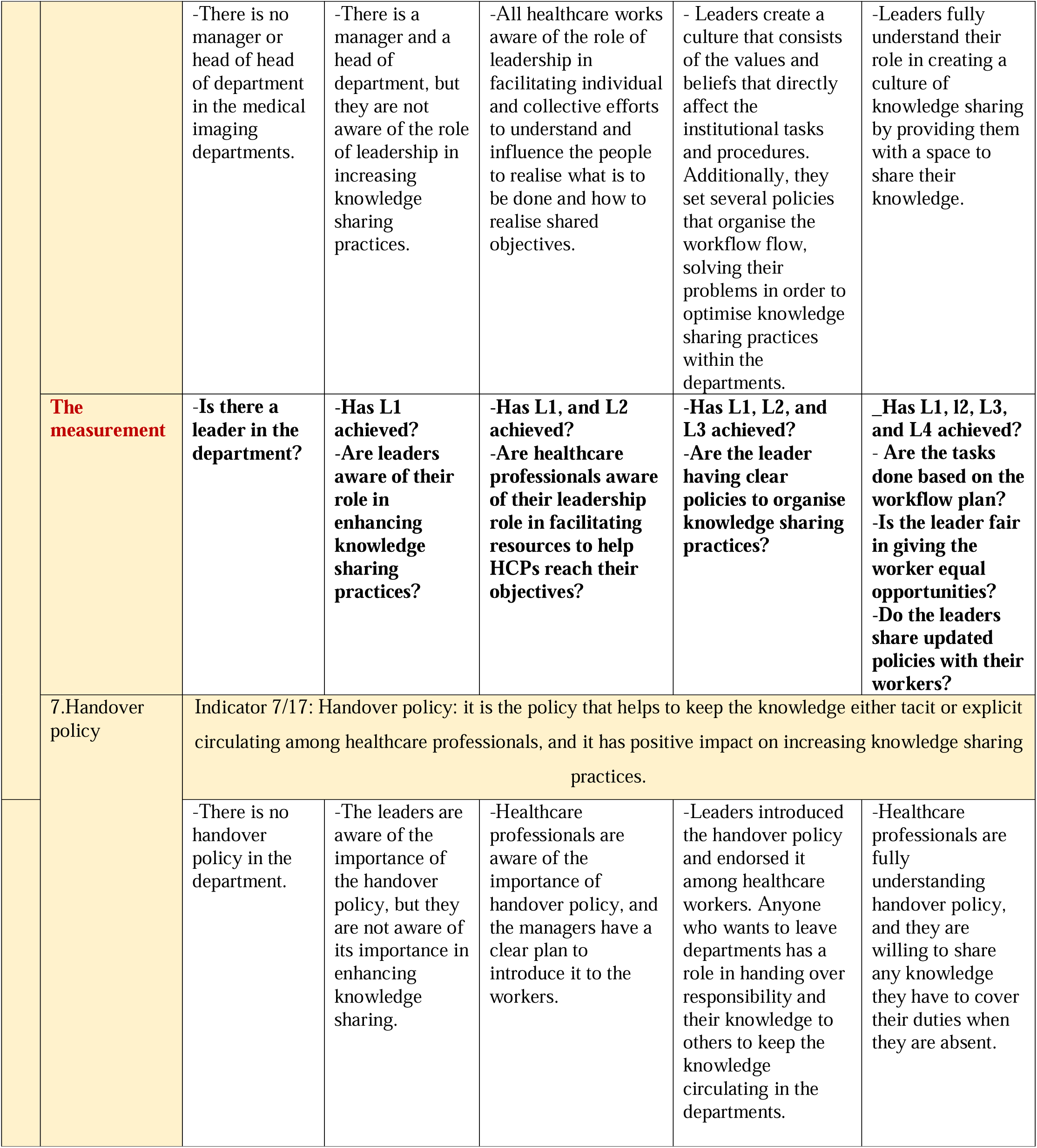
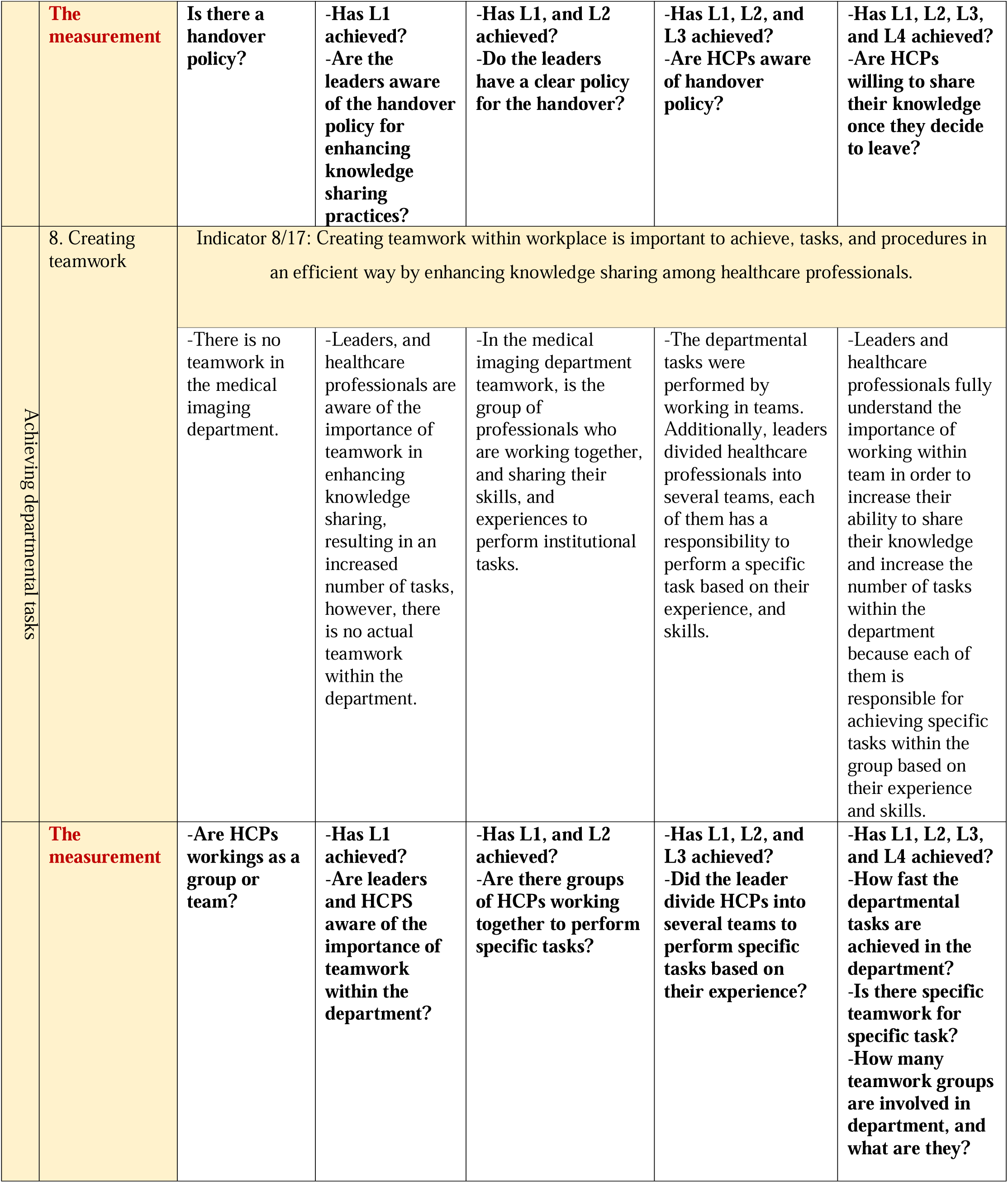
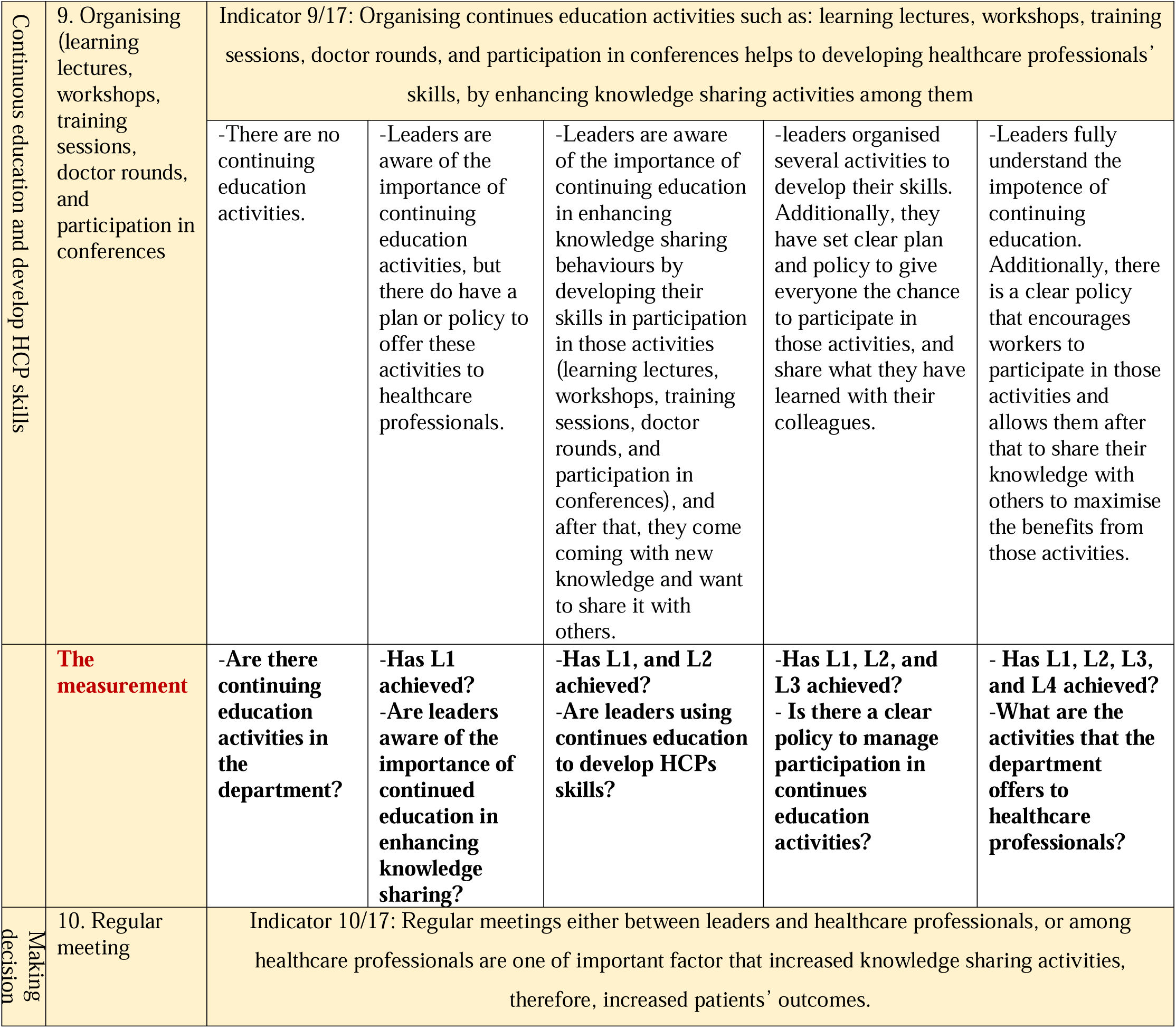
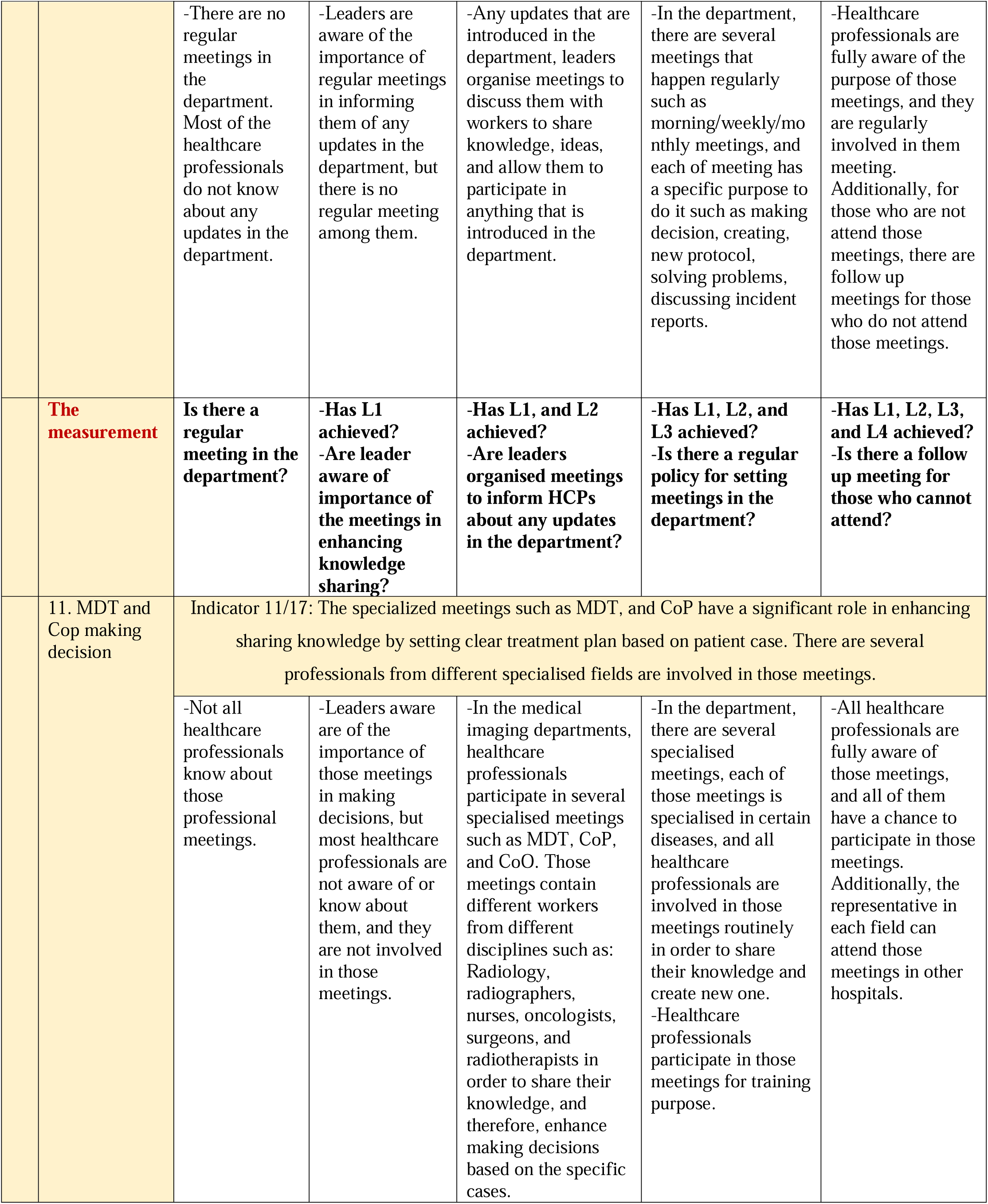
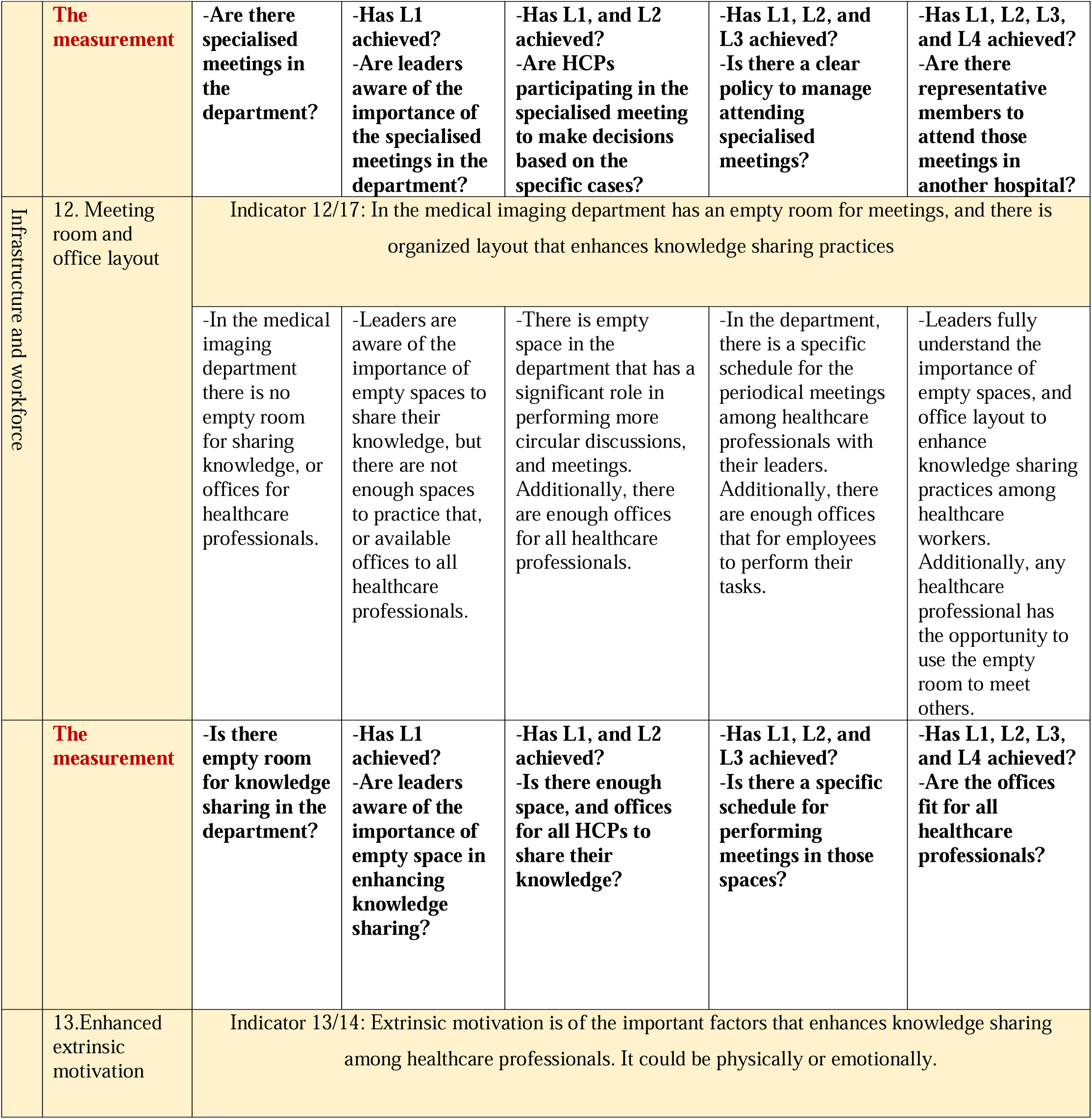
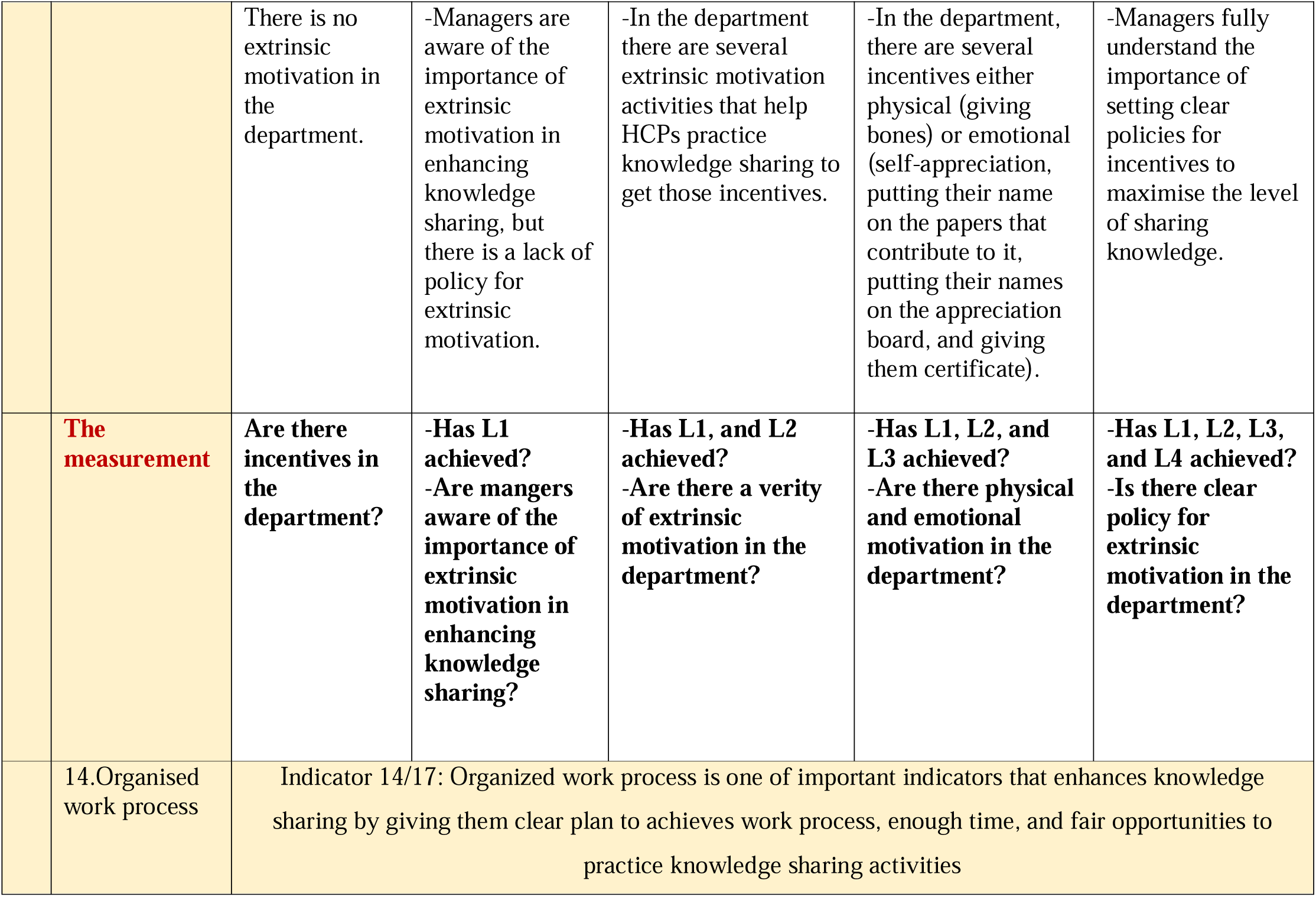
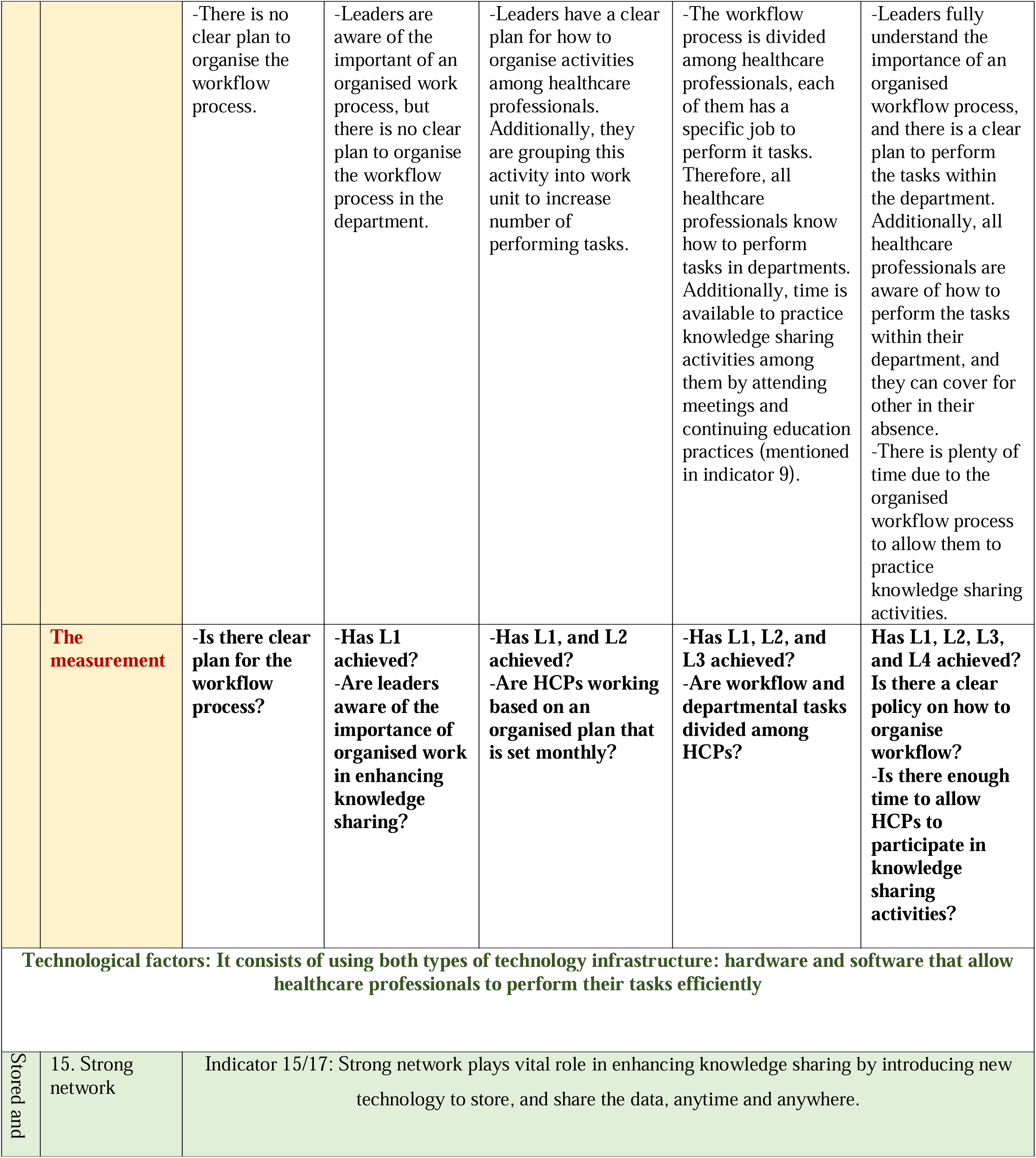
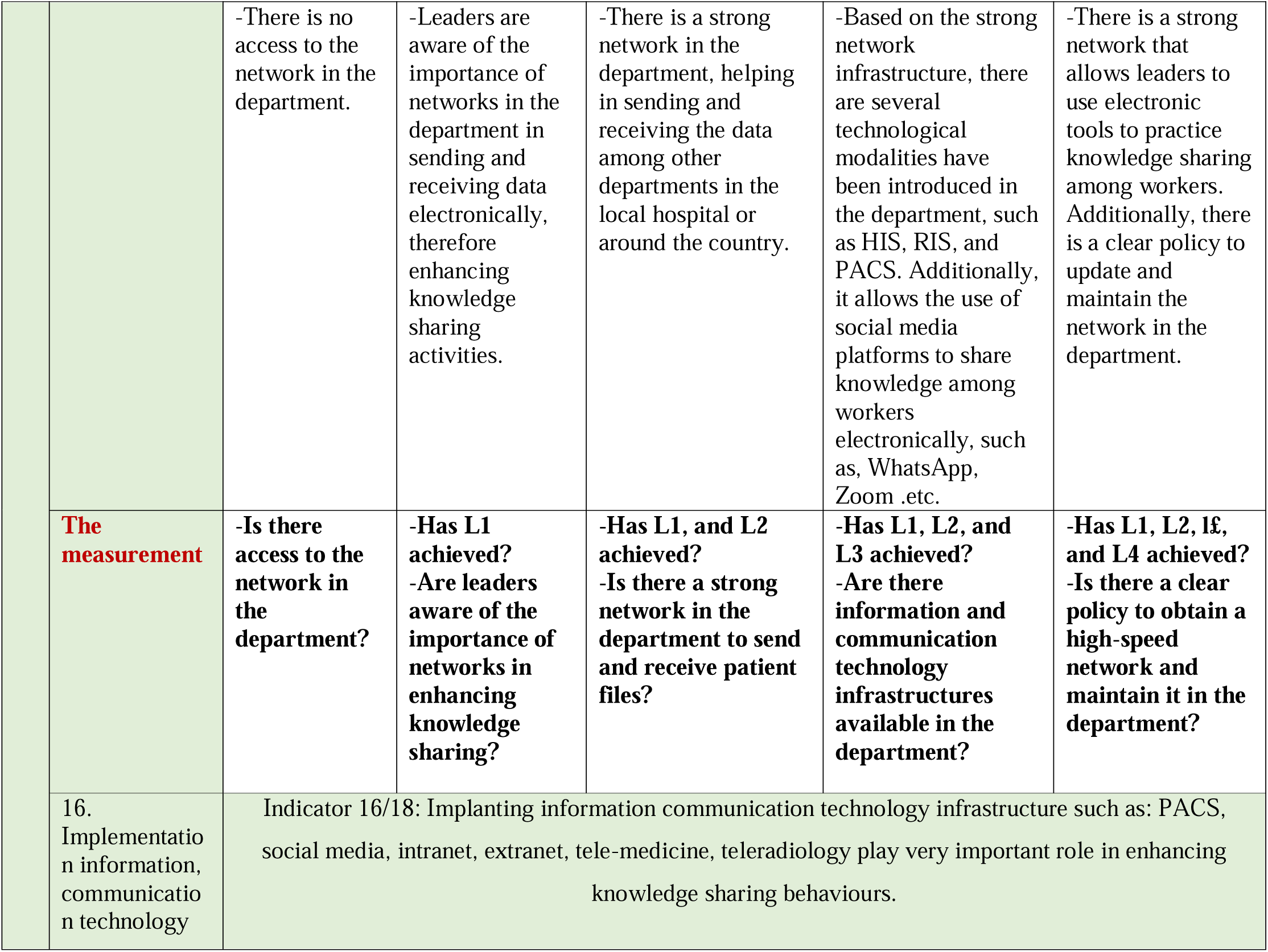
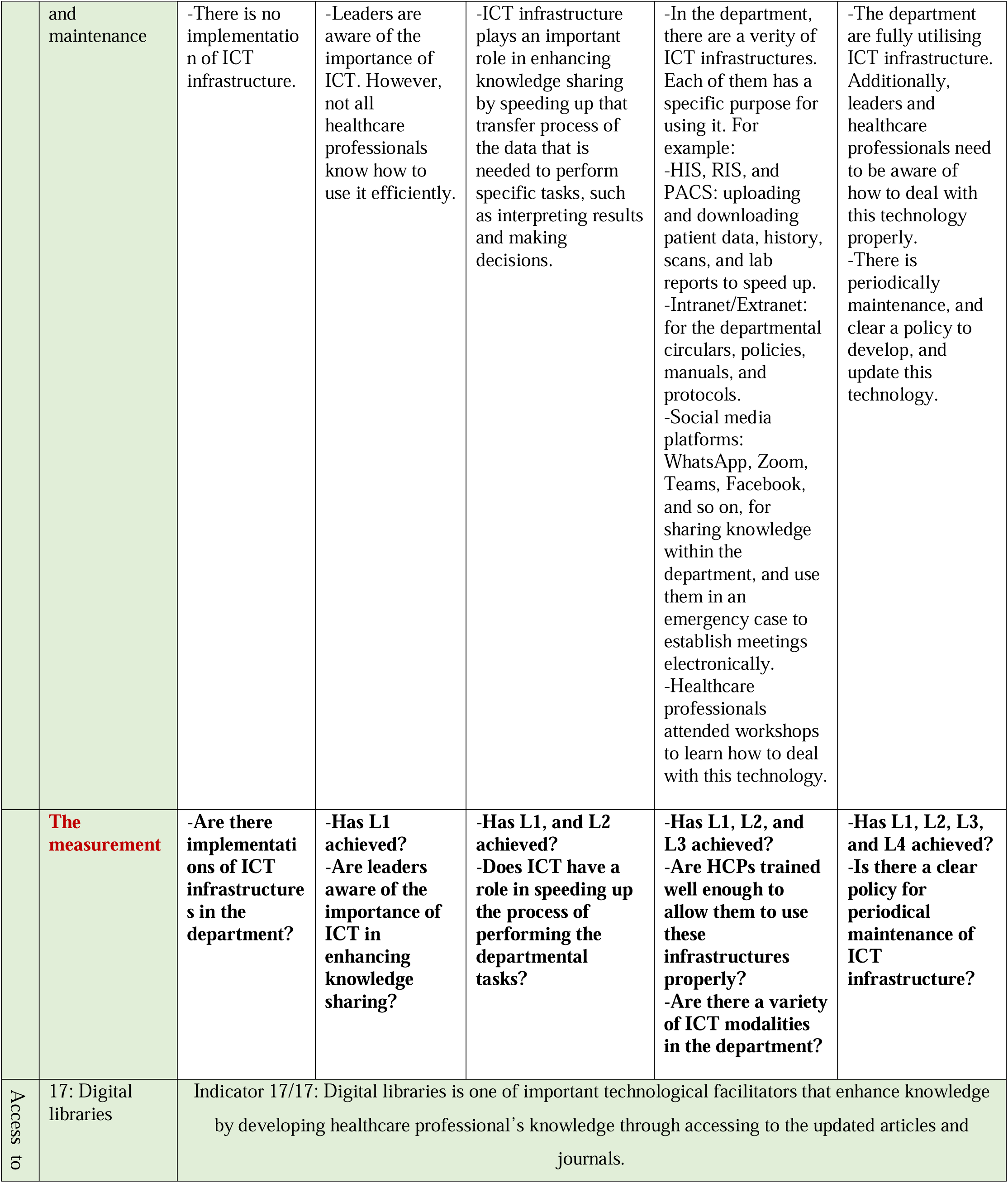
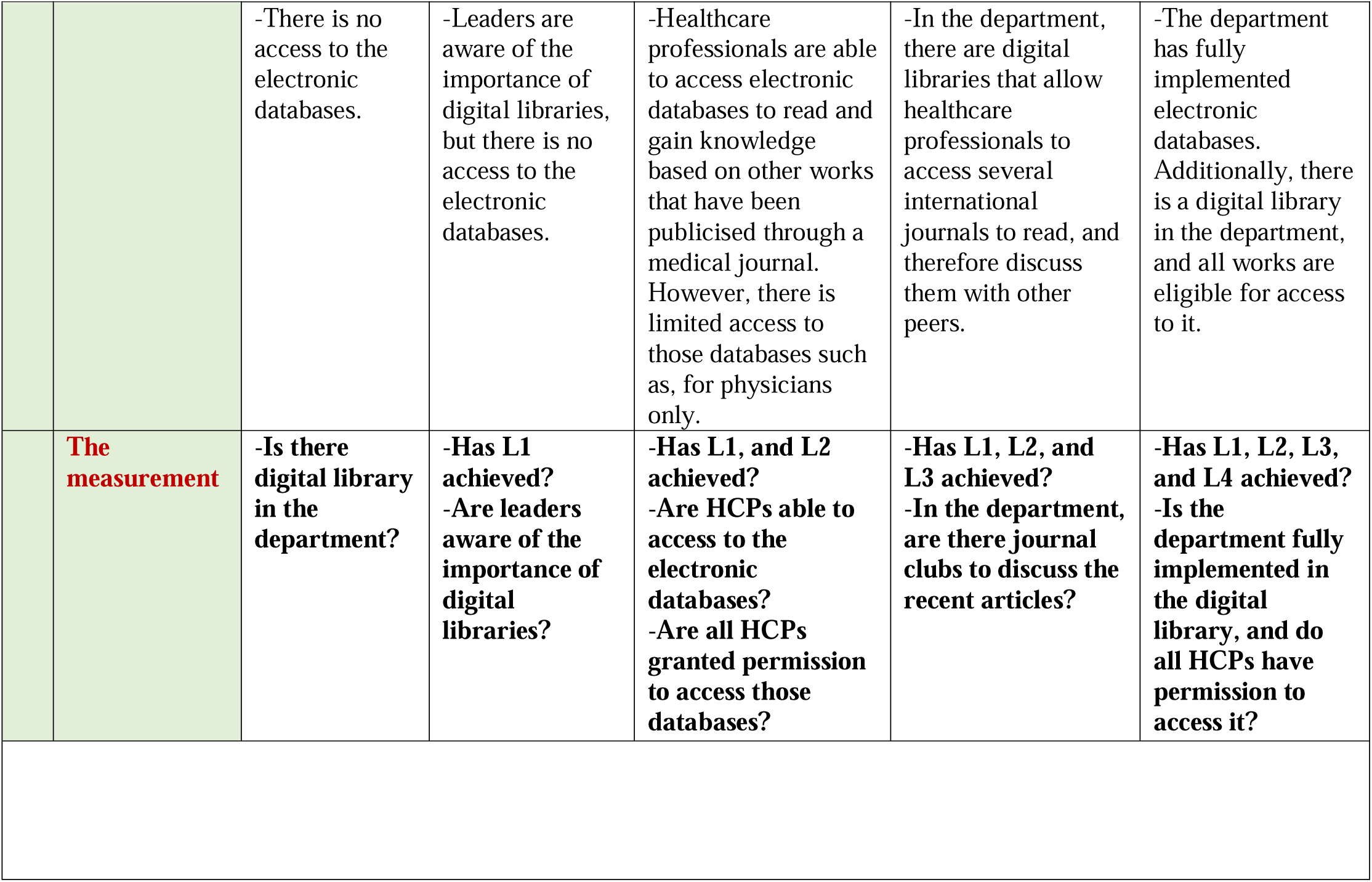
The details of the maturity model for knowledge sharing in medical imaging departments.

## Discussion

Despite the considerable attention to knowledge management in healthcare intuitions, and the significance of the maturity model in managing healthcare recourses, knowledge sharing in hospitals has been less focused on developing the maternity model as a tool for assessing knowledge sharing practices or a roadmap to adopting knowledge sharing behaviours. This is the essence of the present work. This study aimed to develop a knowledge sharing maturity model in the medical imaging department by assessing the factors that affect knowledge sharing practices. Several maturity models have been developed in social media, health systems, digital libraries, ICT, PACS, and telemedicine [39, 41, 43–45]. Those models were used to assess each factor that affects knowledge sharing practices in terms of benchmarking efforts and to develop progressive strategies that might improve its activities. Additionally, Yiren et al., [42] shed light on a maturity model for cancer multidisciplinary teams. That model consists of 17 indicators that are used to measure healthcare professionals’ performance and monitor the quality of performance at cancers centre over time.

Figure 1 highlights the five components that affect KSMM in the medical imaging department and classifies each component in terms of its influence on knowledge sharing behaviours. Awareness of the importance of knowledge management is one of the core components that contribute to the adoption knowledge sharing practices among healthcare professionals. Most of the respondents showed a high level of awareness of the importance of knowledge sharing in developing their skills, increasing healthcare services, and reducing medical errors. Therefore, results showed that more than half of respondents in both cancer centres (17 (58.6%, 32 (57.1%)) participated daily in knowledge sharing activities that were available in their department. Additionally, one of the articles showed that without awareness of the importance of knowledge sharing, there are no knowledge sharing practices in the medical imaging department [10]. The next step is structured types of knowledge (tacit and explicit). Understanding the types of knowledge available in the department, and how to capture, documented, and share it is vital to accelerating knowledge sharing practices in the department. Tacit knowledge appears the in medical imaging department as a dominant type among healthcare professionals that is considered as tool for sharing knowledge in lectures, conferencing, and meetings. In contrast, explicit knowledge takes several forms such as documents, policies, procedures, and manuals. That form allows workers to reach it anytime in an easy way [65]. From the third to the fifth step, factors that affected knowledge sharing that divided into three categories: individual factors, followed by departmental factors, and finally technological factors [10, 38, 47–50, 52, 54–57].

Through this study, the key research questions have been answered. The model can be used as a scoring tool to assess knowledge sharing practices indicators with each maturity model scoring 1, 2, 3, 4, 5 respectively. Moreover, this model can help managers and policymakers find the opportunities for improvements and the way to achieve them. Additionally, applying KSMM helps healthcare institutions increase health services and patient outcomes, reduce errors, and solve the problem in a practical way.

The purpose of using the maturity model in the institutions is not to achieve the highest level of maturity, but the maturity model gives the institutions the roadmap to make a decision about what to improve and how. This study has several limitations that need to be addressed. First, this maturity model was based on the medical imaging department’s vision. In terms of the factors, knowledge sharing factors might from one department to another, but they are similar in general. Therefore, it could be modified to be adopted by other institutions. Second, this study lacks reliability. Therefore, would be useful to apply the Delphi method among experts in a knowledge management in order to evaluate the reliability of this model.

**Figure 2:**
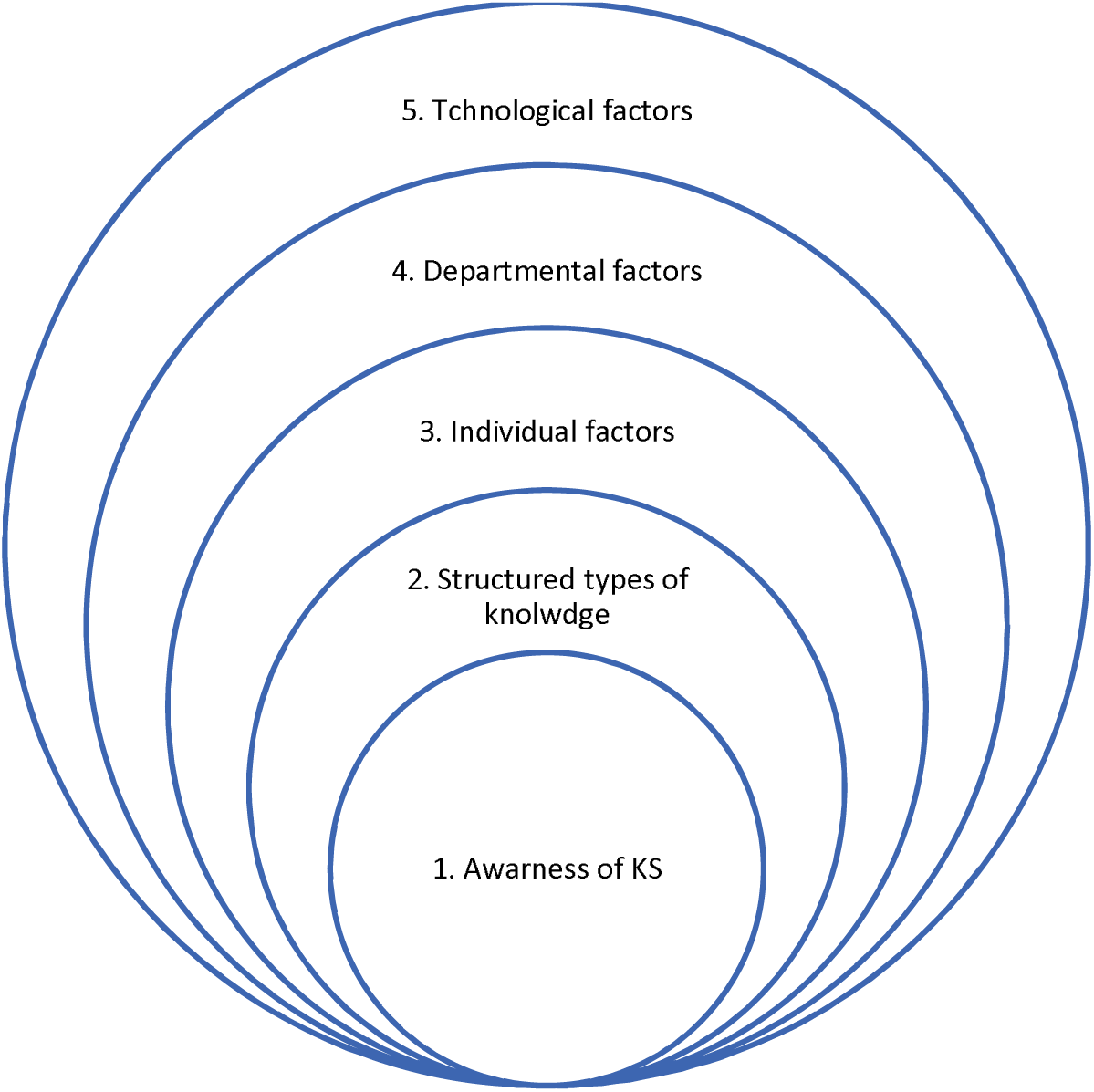
Schematic description of the five components that affect KSMM in medical imaging departments.

## Future work

Following the creation of this knowledge sharing maturity model for medical imaging departments, we will validate this model by applying Delphi methods to provide a consensus view of the suitability of these indicators for improving knowledge sharing practices [66–69]. We will conduct a study in which experts in the field of knowledge management and heads of departments within healthcare organisations will participate in a two round Delphi study assessing the maturity model. This study will extend the results of the work we have presented here, and address the reliability of the model as a generalised tool for use within medical imaging departments and the healthcare sector more widely. We expect that this work will lead to refinements in the model and will provide a practical grounding for the theoretical work presented in this paper.

## Conclusion

Knowledge sharing is considered a core step in the implementation of knowledge management. Health care institutions have a responsibility to adopt knowledge sharing practices in order to manage knowledge that is either tacit or explicit. A medical imaging department is crucial in any healthcare institution. Therefore, creating KSMM is important to develop knowledge-sharing practices. The model proposed in this study allows managers to measure the maturity level of knowledge sharing in the medical imaging department. By providing the roadmap, the knowledge sharing maturity model allows imputations and managers in healthcare intuitions to appraise knowledge sharing practices and adopt a culture of knowledge sharing to achieve the departmental tasks, and improvements. Additionally, it could help managers to assess knowledge sharing practices in the medical imaging department and find the weak points that have a negative on impact those behaviours. A range of factors were addressed from in our previous work and then we evaluated those factors in two medical imaging departments in two cancer centres. Those factors are divided into 5 categories. Therefore, the KSMM consists of 17 indicators that are divided into 11 components and presented in 5 categories. The most important of those indicators is awareness of the importance of knowledge sharing. Presumably because it allows healthcare professionals to develop their skills, and perform several tasks. The model presented might also be used to identify statements for improvement. The measurement of each indicators helps the managers to assess what level they are. If the answer to the first level is NO. Therefore, they have to work on this indicator until they achieve it and next to the next level. Moreover, this model that was presented in table 2 will contribute to identify which they are and assess the weakness points, thereby will help the managers and policy makers to develop knowledge sharing practice to the next level. In general, knowledge sharing practices is important in healthcare institutions to avoid repetitive errors, improve healthcare services, improve collaborations and communication among staff, and therefore it will encourage them to come with new ideas together.

## Supporting information

Multimedia Appendix 2

Multimedia Appendix 1

## Data Availability

All data produced in the present work are contained in the manuscript

## Acknowledgements

This study was completed as a part of doctoral studies funded by Ministry of health, Kuwait, and University of Manchester, UK.

## Multimedia Appendix 1

The questionnaires questions, consent form for the interview, and the questions of the interviews.

## Multimedia Appendix 1

The consent form for the interview, and the questions of the interviews.

## References

1. Kogut B, Zander U. What firms do? Coordination, identity, and learning. Organization science. 1996;7(5):502–18.

2. Mc Evoy PJ, Ragab MAF, Arisha A. The effectiveness of knowledge management in the public sector. Knowledge management research & practice. 2019;17(1):39–51. doi: 10.1080/14778238.2018.1538670.

3. Haughom J. Innovation in healthcare: why it’s needed and where it’s going. Health Catalyst Retrieved from https://www.healthcatalyst.com/innovation-inhealthcare-why-needed-where-going. 2014.

4. Rego I, Pereira L, Dias Á, Gonçalves R, Costa RLd. Knowledge management maturity in healthcare service. International Journal of Knowledge and Learning. 2023;16(1):17–55.

5. Sandhu MS, Jain KK, bte Ahmad IUK. Knowledge sharing among public sector employees: evidence from Malaysia. International Journal of Public Sector Management. 2011;24(3):206–26.

6. Al-Kurdi OF, El-Haddadeh R, Eldabi T. The role of organisational climate in managing knowledge sharing among academics in higher education. Int J Inf Manage. 2020;50:217–27. doi: 10.1016/j.ijinfomgt.2019.05.018.

7. Abzari M, Shahin A, Abasaltian A. Studying the impact of personality constructs on employees’ knowledge sharing behavior through considering the mediating role of intelligent competencies in project-oriented organizations. Modern Applied Science. 2016;10(6):194–204.

8. Vuori V, Okkonen J. Knowledge sharing motivational factors of using an intra-organizational social media platform. Journal of knowledge management. 2012;16(4):592–603.

9. Radević I, Dimovski V, Lojpur A, Colnar S. Quality of healthcare services in focus: the role of knowledge transfer, hierarchical organizational structure and trust. Knowledge Management Research & Practice. 2021:1–12.

10. Al Mashmoum MJ, Hamade SN. Knowledge Sharing Among Employees of Kuwait Cancer Control Center. International Journal of Knowledge Management and Practices. 2019 2019 2021-09-11;7(1):15–25. PMID: 2241046417.

11. Abidi SSR. Knowledge management in healthcare: towards ‘knowledge-driven’decision-support services. International journal of medical informatics. 2001;63(1-2):5–18.

12. Sharma K, Sharma V. Evaluating knowledge management practices in Indian manufacturing and service industry: an overview. International Journal of Knowledge Management Studies. 2018;9(3):222–42.

13. Dorow P, Medeiros C, Camozzato T, Silva C, Vargas F, Huhn A. Knowledge sharing process between radiologist. Int J Adv Res. 2018;6(7):431–6.

14. Klimko G, editor. Knowledge management and maturity models: Building common understanding. Proceedings of the 2nd European conference on knowledge management; 2001: Bled, Slovenia.

15. Serenko A, Bontis N, Hull E. An application of the knowledge management maturity model: the case of credit unions. Knowledge Management Research & Practice. 2016;14(3):338–52.

16. Pee LG, Kankanhalli A. A model of organisational knowledge management maturity based on people, process, and technology. Journal of information & knowledge management. 2009;8(02):79–99.

17. Gibson CF. Managing the four stages of EDP growth. Harvard Business Review. 1974;52(1):76–88.

18. Paulk MC, Curtis B, Chrissis MB, Weber CV. Capability maturity model, version 1.1. IEEE software. 1993;10(4):18–27.

19. Aho M, editor. What is your PMI? A model for assessing the maturity of performance management in organizations. PMA 2012 Conf; 2012.

20. Becker MC, Salvatore P, Zirpoli F. The impact of virtual simulation tools on problem-solving and new product development organization. Research Policy. 2005;34(9):1305–21. doi: 10.1016/j.respol.2005.03.016.

21. Van Aken EM, Letens G, Coleman GD, Farris J, Van Goubergen D. Assessing maturity and effectiveness of enterprise performance measurement systems. International Journal of Productivity and Performance Management. 2005;54(5/6):400–18.

22. Wettstein T, Kueng P. A maturity model for performance measurement systems. WIT Transactions on Information and Communication Technologies. 2002;26.

23. Maier AM, Moultrie J, Clarkson PJ. Assessing organizational capabilities: reviewing and guiding the development of maturity grids. IEEE transactions on engineering management. 2011;59(1):138–59.

24. Jääskeläinen A, Roitto J-M. Designing a model for profiling organizational performance management. International Journal of Productivity and Performance Management. 2015;64(1):5–27.

25. Bititci US, Garengo P, Ates A, Nudurupati SS. Value of maturity models in performance measurement. International journal of production research. 2015;53(10):3062–85.

26. Yan Q-y, Xiang F, Shi X-x, Zhu Q. Implementation of knowledge management in Chinese hospitals. Current medical science. 2018;38(2):372–8.

27. Šajeva S, Jucevičius R. Model of knowledge management system maturity and its approbation in business companies. Socialiniai mokslai. 2010 (3):57–68.

28. Teah HY, Pee LG, Kankanhalli A. Development and application of a general knowledge management maturity model. PACIS 2006 Proceedings. 2006:12.

29. Kulkarni U, St Louis R. Organizational self assessment of knowledge management maturity. 2003.

30. Spanellis A, MacBryde J, Dlllrfler V. A dynamic model of knowledge management in innovative technology companies: A case from the energy sector. European Journal of Operational Research. 2021;292(2):784–97.

31. Carvalho JV, Rocha Á, van de Wetering R, Abreu A. A Maturity model for hospital information systems. Journal of Business Research. 2019;94:388–99.

32. Arif M, Al Zubi M, Gupta AD, Egbu C, Walton RO, Islam R. Knowledge sharing maturity model for Jordanian construction sector. Engineering, Construction and Architectural Management. 2017;24(1):170–88.

33. Mettler T, Rohner P, editors. Situational maturity models as instrumental artifacts for organizational design. Proceedings of the 4th international conference on design science research in information systems and technology; 2009.

34. Lodhi SA. National College of Business Administration & Economics Lahore. 2005.

35. Sayed S, Mutasa R, Kaaya E, Mudenda V, Rajiv E, Vuhahula E, et al. Establishing the College of Pathologists of East, Central and Southern Africa – The Regional East Central and Southern Africa College of Pathology. African Journal of Laboratory Medicine. 2020 2020 2020-06-18;9(1). PMID: 2414179020. doi: 10.4102/ajlm.v9i1.979.

36. Taylor WA, Wright GH. Organizational readiness for successful knowledge sharing: Challenges for public sector managers. Information Resources Management Journal (IRMJ). 2004;17(2):22–37.

37. Supar N, Ibrahim AA, Mohamed ZA, Yahya M, Abdul M, editors. Factors affecting knowledge sharing and its effects on performance: a study of three selected higher academic institutions. International Conference on Knowledge Management (ICKM); 2005.

38. Lema B. A framework to support knowledge sharing practice among health care professionals at Yekatit 12 hospital medical College: Addis Ababa University; 2017.

39. Fitterer R, Rohner P. Towards assessing the networkability of health care providers: a maturity model approach. Information Systems and e-Business Management. 2010;8:309–33.

40. Ekuobase GO, Olutayo VA. Study of Information and Communication Technology (ICT) maturity and value: The relationship. Egyptian Informatics Journal. 2016;17(3):239–49.

41. Sheikhshoaei F, Naghshineh N, Alidousti S, Nakhoda M. Design of a digital library maturity model (DLMM). The Electronic Library. 2018;36(4):607–19.

42. Liu Y, Evans L, Kwan T, Callister J, Poon S, Byth K, et al. Developing a maturity model for cancer multidisciplinary teams. International Journal of Medical Informatics. 2021;156:104610.

43. Van Dyk L, Schutte CS. The telemedicine service maturity model: A framework for the measurement and improvement of telemedicine services. Telemedicine. 2013:217–38.

44. van de Wetering R, Batenburg R. A PACS maturity model: a systematic meta-analytic review on maturation and evolvability of PACS in the hospital enterprise. International journal of medical informatics. 2009;78(2):127–40.

45. Jami Pour M, Jafari SM. Toward a maturity model for the application of social media in healthcare: The health 2.0 roadmap. Online Information Review. 2019;43(3):404–25.

46. Muqadas F, Rehman M, Aslam U, Ur-Rahman U-. Exploring the challenges, trends and issues for knowledge sharing: A study on employees in public sector universities. VINE Journal of Information and Knowledge Management Systems. 2017;47(1):2–15.

47. Monazam Tabrizi N. Models for Describing Knowledge Sharing Practices: The Case Study of UK Hospitals. 2016.

48. Lee HS, Hong SA. Factors affecting hospital employees’ knowledge sharing intention and behavior, and innovation behavior. Osong public health and research perspectives. 2014;5(3):148–55.

49. Shahmoradi L, Safdari R, Piri Z, Mahmodabadi AD, Shahmoradi S, Nejad AF. Knowledge sharing as a powerful base for management: Barriers and solutions. The health care manager. 2017;36(2):176–83.

50. Gider Ö, Ocak S, Top M. Perceptions of physicians about knowledge sharing barriers in Turkish health care system. Journal of medical systems. 2015;39:1–13.

51. Kim S-J, Park M. Leadership, knowledge sharing, and creativity. The Journal of Nursing Administration. 2015;45(12):615–21.

52. Imran MK, Fatima T, Aslam U, Iqbal SMJ. Exploring the Benefits of Social Media Towards Knowledge Sharing Among Doctors. Pakistan Journal of Psychological Research. 2019;34(2).

53. Yip K. Exploring barriers to knowledge sharing: A case study of a virtual community of practice in a Swedish multinational corporation. 2011.

54. Ariati N, Sensuse DI, Handayani PW, editors. Factors affecting knowledge sharing capability of doctors in Palembang. Journal of Physics: Conference Series; 2020: IOP Publishing.

55. Lin TC, Lai MC, Yang SW. Factors influencing physicians’ knowledge sharing on web medical forums. Health Informatics Journal. 2016;22(3):594–607.

56. MamoMulate E, Gojeh LA. Current Status and Factors Affecting Knowledge Sharing Practices among Health Professionals in Hiwot Fana Specialized University Hospital in Ethiopia. 2020.

57. Demsash AW, Chakilu B, Mazengia A. Knowledge Sharing Practice and Its Associated Factors Among Healthcare Providers at University of Gondar Comprehensive Specialized Hospital, North West Ethiopian: Cross-sectional Study. 2021.

58. Almeida MH, Ramos A, Sousa MJ, Santos CM, Fontes AP, editors. Organizational Factors Affecting Knowledge Sharing Capabilities: A Study with Portuguese Health Professionals. European Conference on Knowledge Management; 2020: Academic Conferences International Limited.

59. Pun JK, Matthiessen CM, Murray KA, Slade D. Factors affecting communication in emergency departments: doctors and nurses’ perceptions of communication in a trilingual ED in Hong Kong. International journal of emergency medicine. 2015;8(1):1–12.

60. Almashmoum M, Cunningham J, Alkhaldi O, Anisworth J. Factors That Affect Knowledge-Sharing Behaviors in Medical Imaging Departments in Cancer Centers: Systematic Review. JMIR human factors. 2023;10:e44327-e. doi: 10.2196/44327.

61. Creswell JW, Creswell JD. Research design: Qualitative, quantitative, and mixed methods approaches: Sage publications; 2017. ISBN: 1506386717.

62. Alhalhouli ZT, Hassan Z, Abualkishik AM. An updated model to enhance knowledge sharing among stakeholders in Jordanian hospitals using social networks. Middle-East Journal of Scientific Research. 2013;18(8):1089–98.

63. Noor NM, Salim J. Factors influencing employee knowledge sharing capabilities in electronic government agencies in Malaysia. International Journal of Computer Science Issues (IJCSI). 2011;8(4):106.

64. Lee H-S. Knowledge management enablers and process in hospital organizations. Osong public health and research perspectives. 2017;8(1):26.

65. Adeyelure TS, Kalema BM, Motlanthe BL. An empirical study of knowledge sharing: A case of South African healthcare system. Knowledge Management & E-Learning: An International Journal. 2019;11(1):114–28.

66. Kagias P, Sariannidis N, Garefalakis A, Passas I, Kyriakogkonas P. Validating the Whistleblowing Maturity Model Using the Delphi Method. Administrative Sciences. 2023;13(5):120.

67. Gallotta B, Garza-Reyes JA, Anosike A, editors. Using the Delphi method to verify a framework to implement sustainability initiatives. Proceedings of the International Conference on Industrial Engineering and Operations Management; 2018: 92922.

68. Martins AI, Santinha G, Almeida AM, Ribeiro Ó, Silva T, Rocha N, et al. Consensus on the Terms and Procedures for Planning and Reporting a Usability Evaluation of Health-Related Digital Solutions: Delphi Study and a Resulting Checklist. Journal of Medical Internet Research. 2023;25:e44326.

69. Habibi A, Sarafrazi A, Izadyar S. Delphi technique theoretical framework in qualitative research. The International Journal of Engineering and Science. 2014;3(4):8–13.

