## Supplementary material for "Paper 3: - Towards a Knowledge Sharing Maturity Model for medical imaging departments": Multimedia Appendix 2

**Consent form**

**(Evaluating knowledge sharing practices among employees in the medical imaging department at the Christie hospital and Kuwait Cancer Control Ce**

**If you would like to participate in the interview, please complete and sign the following consent form.**

**•	I………………………..agree to participate in this interview for the research study.**

**•	I understand that even if I choose to participate at this time, I realize that I may withdraw at any moment or refuse to answer any question without any repercussions.**

**•	I consent to having my interview recorded.**

**•	I am aware that any information I offer for this research will be kept confidential.**

**•	I am aware that I am welcome to contact any member of the study team for additional clarity and data.**

**Name of Participant:___________________**

**Signature:____________________**

**Date:__________________________**

**Questions**

**Let me give you a brief information about my research study, the main concern of my research is about evaluating knowledge sharing practices and identifying facilitators that affect knowledge sharing among employees. Sharing knowledge has positive impact on increase patient outcomes and reduce medical error.**

**1.	First of all, tell me about your experience, How long have you been working in the hospital?**

**2. What do you understand of knowledge sharing to be? Give me your definition about KS?**

**3.	In general, do you have a future plan of knowledge sharing ? Would you like to create knowledge sharing culture of the hospital?**

**4.	Are you satisfied with the knowledge sharing approach in your department?**

**5.	Do you believe that knowledge sharing is important among employees in the hospital? Why?**

**6. Do you think knowledge sharing is important in diagnosis cancer cases?**

**7.	In a daily work, what are the current tool available of knowledge sharing in the hospital?**

**8.	Are you satisfying with the current tool for sharing knowledge in your department?**

**9. Do you use online tools to share knowledge in the department?**

**10.	Are you satisfied of using online communicating tools to share knowledge?**

**11.	What are the barriers that hinder knowledge sharing in the workplace?**

**12.	In your opinion, what are challenges of implementing knowledge sharing in the hospital?**

**13.	Tel me more about your department, Did your department encourage and support knowledge sharing among employees?**

**14.	Does the department has an appropriate polices and framework for knowledge sharing among employees?**

**15.	What are the steps that should you or you department take to improve knowledge sharing?**

**For technologist**

**1. First of all, tell me about your experience, How long have you been working in the hospital?**

**2. What do you understand of knowledge sharing to be? Give me your definition about KS?**

**4.	Are you satisfied with the knowledge sharing approach in your department?**

**5.	Do you believe that knowledge sharing is important among employees in the hospital? Why?**

**6. Do you think knowledge sharing is important in diagnosis cancer cases?**

**7.	In a daily work, what are the current tool available of knowledge sharing in the hospital?**

**8.	Are you satisfying with the current tool for sharing knowledge in your department?**

**9. Do you use online tools to share knowledge in the department?**

**10.	Are you satisfied of using online communicating tools to share knowledge?**

**11.	What are the barriers that hinder knowledge sharing in the workplace?**

**12.	In your opinion, what are challenges of implementing knowledge sharing in the hospital?**

**13.	Tel me more about your department, Did your department encourage and support knowledge sharing among employees?**

**14.	Does the department has an appropriate polices and framework for knowledge sharing among employees?**

**15.	What are the steps that should you or you department take to improve knowledge sharing?**
