## Supplementary material for "Paper 3: - Towards a Knowledge Sharing Maturity Model for medical imaging departments": Multimedia Appendix 1

Online Electronic Questionnaire

Dear participants,

Thank you for taking time to fill this questionnaire to assists me with my PhD research study project. I am currently undertaking a Ph.D., the title of which is ''Evaluating knowledge sharing practices among employees in the medical imaging department at the Christie hospital and Kuwait Cancer Control centre''.

The primary aim of this study is to identify the factors that influence knowledge sharing in two hospitals with a comparative analysis of differences between the two organizations. Knowledge sharing in healthcare is defined as “the knowledge sharing in the healthcare institution can be characterized as the dissemination of context-sensitive healthcare knowledge by and for healthcare stakeholders through a collaborative communication medium in order to advance the knowledge quotient of the participating healthcare stakeholders’’ (Abidi 2007, p. 69). There are two types of knowledge, which have been classified by Polyni (1966): Tacit and Explicit. Both types of knowledge will be tested in this study.

The outcome of the study will help policy makers with designing or updating approaches to enhance knowledge sharing practices in those hospitals. In addition, it will help them to face the problems regarding knowledge sharing behaviours and generalise to knowledge sharing across healthcare services.

Data collected will only be used to complete the objectives of the study. You will be asked to fill out an online survey as part of this investigation. You have the option of withdrawing from this study at any time, and your participation is completely voluntary. It takes approximately 10- 15 minutes to complete the survey.

The questionnaire does not ask for any personal information from any of the participants. In the survey, all responses will be kept anonymously. Please keep in mind that there are no right or wrong responses, and that your own opinion is what matters important to us.

Please do not hesitate to contact me if you have any questions or concerns regarding this project, you may send an email to the address below:

Thank you for your participation

**The consent form**:
If you would like to participate in this survey, please consent the following statements.

| 1. I am aware that my participation in this study is voluntary and that I will be free to withdraw at any time without giving a reason and without detriment to myself. |
| --- |
| 2. I understand that any data collected may be included in anonymous form in publication or conference presentations. |
| 3. I am aware that, if it is pertinent to my participation in this research, data acquired during the study may be examined by personnel from The University of Manchester or regulatory authorities. I agree to let these people have access to my data. |
| 4. I understand that a fully anonymised dataset will be deposited in an open data repository. |
| 5. I agree to participate in this study |

**Survey Questions**

**Part1: Demographics profile of respondents**

Please select the most appropriate answer.

1. Which hospital do you work at?
   A. The Christie Hospital
   B. Kuwait Cancer Control Center (KCCC)
2. Which department do you work in?
   A. Brachytherapy & Molecular Radiotherapy Unit
   B. Chemotherapy unit.
   C. Endocrine Unit & PDT
   D. Estates Department
   E. Haematology
   F. Laboratory
   G. Medical imaging department
   H. Medical oncology
   I. Nursing
   J. Nuclear medicine department
   K. Palliative care
   L. Physical therapy
   M. Physics/Radiotherapy
   N. Pharmacy
   O. Radiation oncology
   P. Radiology department
   Q. Radiotherapy unit
   R. Radio pharmacy
   S. Radio pharmacy
   T. Rehabilitation unit
   U. Surgical oncology
   V. Other Technical Services
   W. Other......................................................
3. What is your role?
   A. Administration/clerical
   B. Anaesthetic
   C. Diagnostic Radiographer
   D. Histopathology
   E. Laboratory scientists
   F. Nuclear medicine technologist
   G. Nurse
   H. Pharmacists
   I. Radiology nurses
   J. Specialist doctor (Consultants)
   K. Others ..............................................
4. What is your sex?
   A. Male
   B. Female
   C. Prefer not to say
   D. None of the above
5. What is your age group?
A. Under 20 years
B. 20—30 years.
C. 30-40 years.
D. 40-50 years.
E. 50-60 years.
F. Above 60 years.
6. What is your highest education level?
   A. Diploma
   B. First degree (Bachelor)
   C. Master’s degree
   D. Doctorate degree
   E. Other...............................................
7. How many years you have been in this hospital?
   A. Less than 10 years
   B. 10-20 years
   C. 20-30 years
   D. More than 30 years

**Part 2: Questions about knowledge sharing background**

Knowledge sharing is part of the knowledge management cycle. It is defined as sharing ideas, thoughts, and experiences among employees, for example in morning meeting sessions, multidisciplinary team meetings (MDTs), Conferences, etc. These practices can improve patients’ outcomes and minimize medical errors. In addition, it can help make such shared knowledge reusable for all employees.

Based on the above knowledge sharing definition please answer the following question.

8. In the daily workplace, how often do you share knowledge with other staff, for example, multi-disciplinary team meetings (MDTs), morning sessions. etc?
   A. Daily
   B. Weekly
   C. Monthly
   D. Every six month
   E. Never
9. In your current role what mechanisms do you use to share knowledge? (Possible to choose more than one answer)
   A. Emails
   B. Face to Face communication.
   C. Microsoft Teams
   D. Observation
   E. PACS (Picture Achieving and Communication System).
   F. Using social media (e.g., What’s app, Facebook, twitter, so on)
   G. Using phone calls
   H. Other..............................
10. How motivated are you to try and share knowledge with others?
    A. Very low
    B. Low
    C. Medium
    D. High
    E. Very high
11. In your hospital, are the incentives or polices in place to encourage knowledge sharing?
    A. Yes
    B. No

**For the following questions please tick (√) one answer only which indicates your extent of agreement: This section examines facilitators of Knowledge sharing. These facilitators are classified into three categories: individuals, departmental, and technological facilitators.**

| Knowledge sharing facilitators | | | | | | | | |
| --- | --- | --- | --- | --- | --- | --- | --- | --- |
| Individuals Facilitators | | **Strongly disagree** | **Disagree** | **Somewhat disagree** | **Neither agree nor disagree** | **Somewhat agree** | **Agree** | **Strongly agree** |
| 10. | **In the hospital, there are periodic meetings in which employees**  **working in different disciplines, may participate.** |  |  |  |  |  |  |  |
| 11. | **In the hospital, there are continuous education programmes such as training courses and workshops within hospital, in which employees can participate.** |  |  |  |  |  |  |  |
|  | **Trust** |  |  |  |  |  |  |  |
| 12. | **I feel fully confident in my own knowledge, and I want to share it with others.** |  |  |  |  |  |  |  |
| 13. | **I trust in knowledge of my colleagues and information that they shared with me.** |  |  |  |  |  |  |  |
| 14. | **If I share my knowledge, my colleagues will feel confident about my ideas, skills, and capabilities to enhance knowledge sharing.** |  |  |  |  |  |  |  |
| 15. | **If I faced problems during work, my colleagues would try to help me resolve them.** |  |  |  |  |  |  |  |
|  | **Awareness** |  |  |  |  |  |  |  |
| 17. | **I am aware of the importance of knowledge sharing among employees at workplace** |  |  |  |  |  |  |  |
| 18. | **Knowledge sharing among employees helps to prevent mistakes that could happen during day-to-day work.** |  |  |  |  |  |  |  |
| 19. | **Sharing knowledge with my colleagues will help me to gain new skills, ideas.** |  |  |  |  |  |  |  |
| 20. | **Knowledge sharing behaviours help employees to learn faster** |  |  |  |  |  |  |  |
|  | **Positive attitudes** |  |  |  |  |  |  |  |
| 21. | **I believe that positive attitudes will help to enhance knowledge sharing among others** |  |  |  |  |  |  |  |
| 22. | **Positive attitudes have a significant role in increasing knowledge sharing behaviours** |  |  |  |  |  |  |  |
| 23. | **I believe that positive attitudes are the first step to sharing knowledge** |  |  |  |  |  |  |  |
|  | **Experience** |  |  |  |  |  |  |  |
| 24. | **I have a good amount of experience that I can share it with my colleagues.** |  |  |  |  |  |  |  |
| 25. | **I believe that experience plays a significant role in sharing knowledge** |  |  |  |  |  |  |  |
| 26. | **Knowledge sharing behaviours will increase when employees have enough experience.** |  |  |  |  |  |  |  |
|  | **Personality** |  |  |  |  |  |  |  |
| 27. | **I have confidence in my ability, to share knowledge.** |  |  |  |  |  |  |  |
| 28. | **I enjoy sharing my knowledge with colleagues.** |  |  |  |  |  |  |  |
| 29. | **I am open minded and receptive to new ideas.** |  |  |  |  |  |  |  |
|  | **Self-esteem** |  |  |  |  |  |  |  |
| 30. | **I believe that self-esteem is an important aspect in sharing knowledge** |  |  |  |  |  |  |  |
| 31. | **I have confidence in my ability to successfully share knowledge with colleagues.** |  |  |  |  |  |  |  |
|  | **Self-efficacy** |  |  |  |  |  |  |  |
| 32. | **I believe that self-efficacy is important to motivate us to share knowledge** |  |  |  |  |  |  |  |
| 33. | **I have the self-efficacy to share my knowledge with others** |  |  |  |  |  |  |  |
|  | **Intrinsic- motivation** |  |  |  |  |  |  |  |
| 34. | **I believe that I have knowledge that will help in increasing productivity.** |  |  |  |  |  |  |  |
| 35. | **I feel happy when I am helping my colleagues by sharing my knowledge with them** |  |  |  |  |  |  |  |
| Departmental facilitators | | | | | | |  |  |
|  | **Extrinsic motivation** |  |  |  |  |  |  |  |
| 36. | **There is acknowledgement for employees who share their knowledge from the hospital** |  |  |  |  |  |  |  |
| 37. | **Sharing knowledge will help me to advance in my career.** |  |  |  |  |  |  |  |
|  | **Leadership: leadership is an employee who have responsibilities to manage work among several employees in one department such as: head of department, and senior management.** | | | | | | | |
| 38. | **I believe that the hospital leadership has a responsibility to encourage and improve knowledge sharing activity.** |  |  |  |  |  |  |  |
| 39. | **I believe that leaderships play an important role in minimizing conflict** |  |  |  |  |  |  |  |
| 40. | **The head of department or senior management has a positive impact on enhancing knowledge sharing.** |  |  |  |  |  |  |  |
|  | **Teamwork** |  |  |  |  |  |  |  |
| 41. | **I believe that teamwork has a significant role in sharing knowledge** |  |  |  |  |  |  |  |
| 42. | **Teamwork is a part of daily work in each department that enhance knowledge sharing.** |  |  |  |  |  |  |  |
| 43. | **Teamwork has a positive impact on enhancing well-being among employees.** |  |  |  |  |  |  |  |
|  | **Culture** |  |  |  |  |  |  |  |
| 44. | **I believe that a culture of communicating is important to enhance knowledge sharing.** |  |  |  |  |  |  |  |
| 45. | **Cultural collaboration has a significant role in sharing knowledge among employees.** |  |  |  |  |  |  |  |
| Communities of Practice are defined as types of meeting or working groups that takes place among members of a healthcare community from within different fields in order to share knowledge. | | | | | | | | |
| 45. | **There are communities of practice in the hospital that I can you for knowledge sharing.** |  |  |  |  |  |  |  |
| 46. | **I believe that communities of practice play a significant role in enhancing knowledge sharing among employees** |  |  |  |  |  |  |  |
| 47. | **Multidisciplinary team meetings are important to increase patient’s outcomes and reduce errors.** |  |  |  |  |  |  |  |
|  | **Learning and training** |  |  |  |  |  |  |  |
| 48. | **I believe that workshops have a significant impact on knowledge sharing** |  |  |  |  |  |  |  |
| 49. | **In the hospital, there are workshops and training sessions that enhance my learning and knowledge sharing.** |  |  |  |  |  |  |  |
| 50. | **The hospital encourages employees to participate in conferences locally and internationally.** |  |  |  |  |  |  |  |
| 51. | **I believe that morning meeting sessions have a positive impact on knowledge sharing.** |  |  |  |  |  |  |  |
|  | **Departmental arrangements** |  |  |  |  |  |  |  |
| 52. | **In the hospital, there is a conference room or meeting room can be used for knowledge sharing.** |  |  |  |  |  |  |  |
| 53. | **I believe that offering an open space to share knowledge is part of a department’s responsibility.** |  |  |  |  |  |  |  |
| 54. | **In the hospital, knowledge sharing practices are part of daily working practice.** |  |  |  |  |  |  |  |
|  | **Doctor rounds** |  |  |  |  |  |  |  |
| 55. | **I believe that daily doctor rounds are an important way of improving knowledge.** |  |  |  |  |  |  |  |
| 56. | **In the hospital, there are daily rounds for professional employees to help develop skills.** |  |  |  |  |  |  |  |
| Technological facilitators | | | | | | |  |  |
|  | **ICT (Information communication technology)** |  |  |  |  |  |  |  |
| 57. | **In the hospital, there is information communication technology infrastructure (e.g., Intranet, Extranet, PACS, and so on)** |  |  |  |  |  |  |  |
| 58. | **I believe that social media has a significant impact on knowledge sharing behaviours** |  |  |  |  |  |  |  |
| 59. | **There are technical support and maintenance groups available to address information communication technology related problems.** |  |  |  |  |  |  |  |
| 60. | **Employees in the hospital have the knowledge and skills to use information communication technology effectively.** |  |  |  |  |  |  |  |
| 61. | **Employees in the hospital use social information communication technology to communicate with each other.** |  |  |  |  |  |  |  |
|  | **Network** |  |  |  |  |  |  |  |
| 62. | **In the hospital, there is a high-speed network available.** |  |  |  |  |  |  |  |
| 63. | **I believe that an available network is vital in enabling knowledge sharing.** |  |  |  |  |  |  |  |
|  | **Digital library: It is an electronic recourse such as eBooks and databases, which are related to the medical background and published articles that support making decisions.** | | | | | | | |
| 64. | **I believe that digital libraries facilitate learning and therefore knowledge sharing.** |  |  |  |  |  |  |  |

**65. Feel Free to add any Comment regarding Knowledge sharing practices**

**Thank you**
